# Ecological characterisation of frequency-domain local field potential biomarkers in essential tremor

**DOI:** 10.64898/2026.11.25.26368820

**Authors:** Edouard Courtin, Sahar Allouch, Margot Bakes, Claire Brandet, Olivier Branchard, Mathilde Guibourg De Luzinais, Emmanuel Cuny, Pierre Burbaud, Mahmoud Hassan, Dominique Guehl

## Abstract

Adaptive deep brain stimulation could make the treatment of essential tremor more responsive to patients’ needs, delivering stimulation when tremor-provoking movements occur rather than continuously, thereby potentially reducing habituation-related loss of efficacy and stimulation-induced adverse effects. Its success, however, depends on neural control signals that remain measurable and meaningful long after surgery, at home rather than in the laboratory, and while stimulation is running. Thalamic local field potentials offer such candidate signals, yet they have largely been characterised intraoperatively or in the early postoperative period, under supervised conditions that reveal little about their behaviour in daily life. We therefore followed nine patients with essential tremor implanted with a sensing-enabled neurostimulator targeting the ventral intermediate nucleus and posterior subthalamic area, combining in-clinic recordings three and six months after surgery with weekly, patient-performed home recordings. We extracted 13 spectral features encompassing canonical and tremor-frequency band power, aperiodic spectral properties and periodic beta activity, and evaluated their agreement between home and clinic recordings, between-session reliability and movement-related modulation across recording settings and stimulation states. Oscillatory features did not differ systematically between home and clinic, whereas the aperiodic background was consistently steeper at home and poorly reproducible. Periodic beta measures emerged as the most robust candidates: with stimulation off, their clinic–home reliability approached that observed between the two clinic visits, and a postural task suppressed beta activity across recording settings, an effect carried by the periodic component rather than by the aperiodic background. Stimulation, however, reduced the reliability of beta measures and markedly attenuated their movement-related modulation, precisely under the conditions in which an adaptive controller would need to operate. Within the limits of this small cohort, thalamic beta activity therefore appears unlikely to serve as a stand-alone control signal for adaptive stimulation in essential tremor, highlighting the potential need for complementary cortical or peripheral inputs.

## Introduction

Essential tremor is one of the most common movement disorders in adults, affecting approximately 1% of the general population, with prevalence increasing markedly with age.^1^ It is characterised by a progressive action tremor predominantly affecting the upper limbs, with substantial inter-individual variability in severity and clinical course.^2^ Although several pharmacological treatments are available, many patients remain refractory to medication and experience considerable impairment in activities of daily living.^3,4^ For these patients, deep brain stimulation (DBS) of the ventral intermediate nucleus (VIM) of the thalamus or the posterior subthalamic area (PSA) has been an established surgical treatment for decades^5^ providing substantial and sustained tremor reduction.^6^

Maintaining long-term efficacy, however, remains a major challenge in thalamic DBS for essential tremor. A substantial proportion of patients experience a decline in stimulation benefit over years of follow-up,^7–9^ a phenomenon thought to reflect, at least in part, neural adaptation to chronic thalamic stimulation.^8^ Loss of efficacy is commonly managed by escalating stimulation parameters, which increases exposure to adverse effects such as dysarthria and gait disturbance.^10^ More fundamentally, this long-term limitation may relate to how stimulation is delivered. Conventional DBS is applied continuously, irrespective of tremor state, whereas the clinical expression of essential tremor is strongly state-dependent, with disability occurring predominantly during voluntary movement and sustained posture.^2,11^ This mismatch provides a strong physiological rationale for closed-loop, demand-driven stimulation that delivers therapy selectively during tremor-provoking motor states, an approach already shown to be feasible in essential tremor.^11,12^

Implementing such systems requires reliable neural, state-dependent biomarkers that can, ideally, be measured directly from the implanted electrodes. Local field potentials (LFPs) recorded at the stimulation target provide access to oscillatory activity within the cerebello-thalamo-cortical circuit implicated in tremor generation.^13,14^ Several candidate control signals have been identified in the VIM, including oscillatory activity at tremor frequency^15–18^ and movement-related suppression of beta-band activity.^19,20^ Proof-of-concept studies in which thalamic local field potentials triggered stimulation during tremor-provoking movements achieved tremor control comparable to continuous stimulation, albeit acutely, within days of implantation;^11,21^ chronic embedded closed-loop systems in essential tremor have so far relied on cortical sensing.^12,22^

A major limitation, however, is that most characterisations of VIM local field potential biomarkers have been either performed intraoperatively or during the early postoperative period. Recordings obtained shortly after electrode implantation may be influenced by the microlesion effect, which can suppress tremor for weeks to months after thalamic surgery^6,23^ and transiently alters local field potential dynamics, as shown in the subthalamic nucleus.^24^ More importantly, biomarkers intended for long-term adaptive DBS (aDBS) must remain detectable and physiologically meaningful months after implantation and under the less controlled conditions of everyday life.^22,25^ Chronic and repeated recordings have been investigated in the subthalamic nucleus in Parkinson’s disease,^26,27^ whereas longitudinal thalamic recordings in essential tremor remain confined to small cohorts studied in the laboratory.^19,22^ In particular, it is not known whether candidate VIM/PSA spectral biomarkers measured in the clinic can be reproduced reliably during repeated, longitudinal ecological recordings at home. Bidirectional neurostimulators capable of chronic, home-based spectral recording^28,29^ now make it possible to address this question directly.

In this study, we characterised VIM/PSA LFP features across two in-clinic assessments and repeated, patient-performed home recordings acquired weekly in the intervening period, first to evaluate their reproducibility across sessions, recording environments and stimulation states, and second to characterise their movement-related modulation.

## Materials and methods

### Participants and study design

We prospectively studied ten patients with severe, disabling, medication-refractory essential tremor treated with bilateral DBS of the ventral intermediate nucleus/posterior subthalamic area (VIM/PSA) using the Percept bidirectional neurostimulator (Medtronic, Minneapolis, MN) as part of standard clinical care. All participants fulfilled the 2018 consensus criteria of the International Parkinson and Movement Disorder Society.^2^ Full inclusion and exclusion criteria, as well as detailed surgical procedure are provided in Supplementary Methods S1 and S2. The study was approved by the Comité de Protection des Personnes Nord Ouest IV (Lille, France; no. 22.04558.000153), and all participants provided written informed consent.

Participants were studied over a three-month period, from three (M3) to six months (M6) after implantation, with two in-hospital supervised recording sessions at M3 and M6 and weekly patient-performed home-based “snapshot” recordings between visits (Fig. 1A).

Tremor severity was assessed at M3 using the Fahn–Tolosa–Marin Rating Scale and compared with the preoperative assessment to quantify DBS-associated clinical improvement.

Stimulation settings were largely unchanged between M3 and M6 (Supplementary Table 2). Stimulation contacts, pulse width and frequency remained unchanged in all patients except P02, in whom the left directional contact was changed between sessions. Per-contact current was self-titrated between visits by less than 0.5 mA in three patients (P01, P02, P04) to optimise clinical outcome.

**Figure 1.**
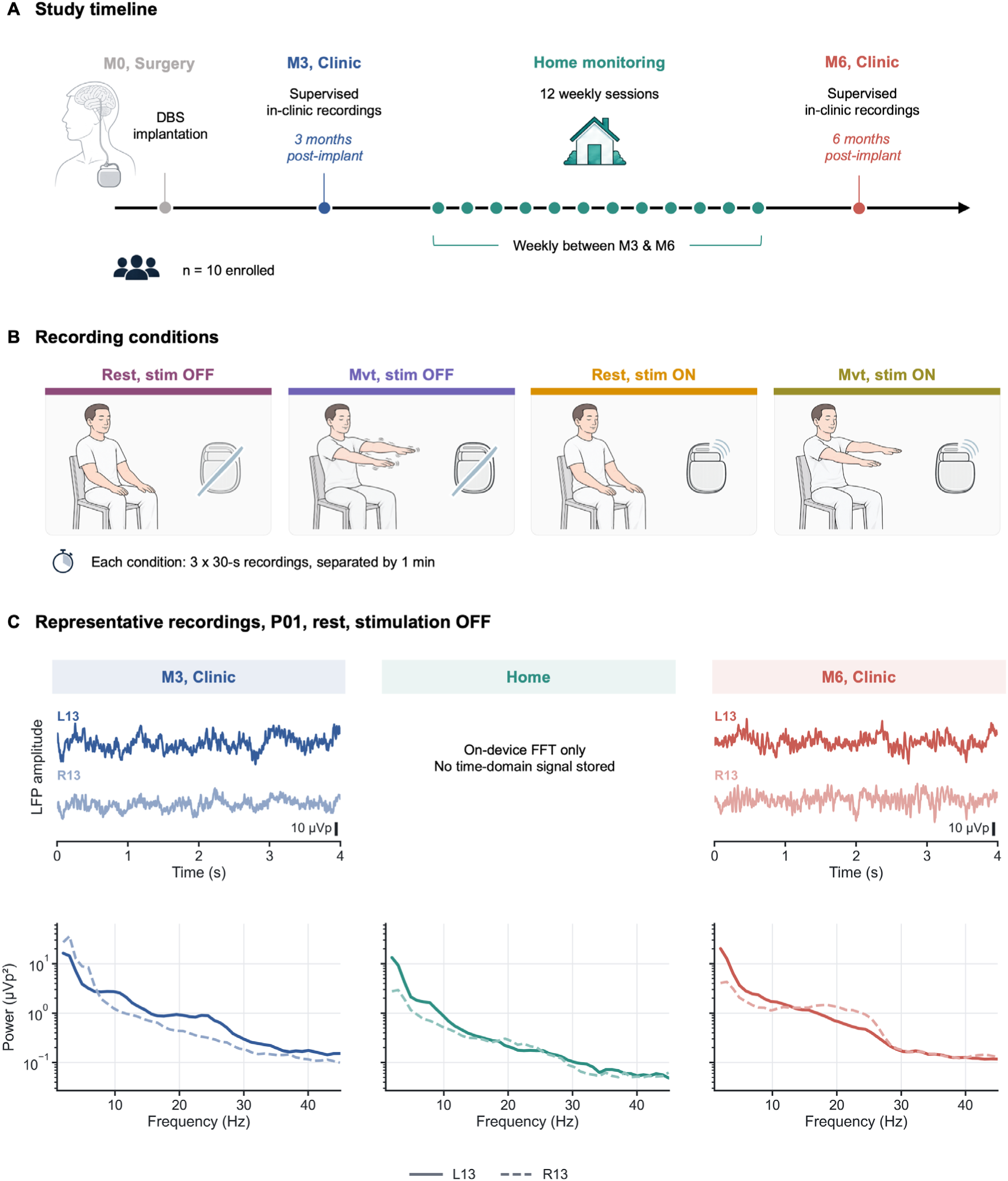
Study design and representative thalamic LFP recordings. **(A)** Study timeline showing DBS implantation, supervised clinic recordings at 3 (M3) and 6 months (M6) post-implantation, and weekly home recordings between visits. **(B)** Recording conditions comprised rest and a tremor-provoking task with stimulation OFF and ON; each condition included three 30-s recordings separated by 1 min. **(C)** Representative bilateral LFP time series and power spectra from participant P01 during rest with stimulation OFF recorded from the bipolar channel between contact 1 and 3 on the left (L13) and right (R13) leads. Home recordings comprised on-device FFT spectra only; time-domain signals were not stored. DBS = deep brain stimulation; FFT = fast Fourier transform; LFP = local field potential.

### LFP acquisition in clinical and home settings

Local field potentials were recorded from the DBS electrodes in two complementary settings and under four conditions: rest and a tremor-provoking postural task, each with stimulation OFF and ON (Fig. 1B).

#### In-hospital

Recordings were performed at M3 and M6 using two “sensing” modes. With stimulation OFF, time-domain signals were streamed simultaneously from all available bipolar contact pairs (“Indefinite streaming”) after a 5-min stimulation washout (Fig. 1C). With stimulation ON, sensing was restricted to a single bipolar configuration comprising the two contacts flanking the active stimulation contact, allowing recording during ongoing stimulation (“BrainSense streaming”). This sensing-friendly bipolar montage was subsequently used for the home recordings and for all analyses. Neural signals were recorded simultaneously with accelerometry positioned on the dorsal aspect of the index, surface EMG and ECG, and a brief transcutaneous electrical pulse was delivered to synchronise neural and peripheral signals.

#### Home setting

Patients performed standardised home recordings once weekly between M3 to M6 using the sensing-friendly configuration defined above. For each of the four conditions, three 30-s recordings were acquired and stored as device “event” snapshot. Home recordings therefore comprised device-derived spectral data rather than stored time-domain signals (Fig. 1C). Stimulation-OFF recordings were obtained upon waking, following an overnight stimulation washout, to minimise carry-over effects. Recordings were initiated by an accompanying person using the handheld controller, and patients documented any deviations from the protocol. At M6, patients rated the ease of performing the home procedure on a 10-point visual analogue scale (0 = very difficult, 10 = very easy).

### LFP preprocessing and spectral estimation

Clinical recordings acquired at M3 and M6 consisted of raw LFP time series sampled at 250 Hz. To maintain comparability with the minimally processed spectra generated on-chip by the implanted device, no band-pass or notch filtering and no automated artefact correction were applied. Recordings were visually inspected for transcutaneous electrical nerve stimulation (TENS) synchronisation pulses, high-amplitude transients and other technical artefacts, and segments containing clearly identifiable artefacts were excluded.

Home event snapshots contained spectra computed on-chip by the implanted neurostimulator. These were expressed as single-sided fast Fourier transform (FFT) amplitudes in microvolt-peak per frequency bin (µVp), rather than as conventional power spectral density in µV²/Hz. Clinical spectra were therefore estimated offline using parameters chosen to reproduce the device output. For each channel, Welch’s method was applied using 1-s Hann windows (250 samples), 80% overlap and a 256-point FFT, with zero-padding from 250 to 256 samples. The resulting spectral density, *Pxx* (µV²/Hz), was converted to microvolt-peak amplitude as √*Pxx* × 1.695, where the scaling factor accounts for the Hann-window root mean square (RMS), peak-to-RMS conversion, single-sided scaling and zero-padding.

All subsequent analyses were performed in the power domain. Spectral amplitudes in µVp were squared to obtain power-like values in µVp². For clinical recordings, Welch’s method yielded a single mean spectrum per channel. For home recordings, the corresponding µVp² spectra were averaged across repeated snapshots.

### Spectral feature extraction

Given the limited availability of spectral data from home recordings, analyses requiring access to the underlying time series, such as burst detection, phase-based coupling and time-resolved spectral estimation, could not be performed consistently across settings. To ensure direct comparability between clinical and home recordings, we therefore focused on features that could be extracted from the available spectra in both settings. Three complementary families of spectral features were considered. First, power within canonical frequency bands was quantified to enable comparison with previous thalamic LFP studies. Second, given the interindividual variability in tremor frequency and its proximity to canonical band boundaries, power was estimated within patient-specific bands centred on the tremor frequency and its second harmonic. Third, because changes in band power may reflect variations in the broadband aperiodic background rather than oscillatory activity alone, spectra were parameterised to distinguish periodic and aperiodic components. This allowed the extraction of additional candidate biomarkers and band-power measures adjusted for the aperiodic background.

#### Canonical band power

Absolute band power was computed by summing the squared values over all frequency bins falling within each predefined band: delta (1–4 Hz), theta (4–8 Hz), alpha (8–13 Hz) and beta (13–35 Hz). For each participant, session, condition and hemisphere, this procedure yielded a single band-power estimate for each predefined frequency band. Gamma power (>35 Hz) was excluded because of high-frequency censoring (see Supplementary Methods S8 and Supplementary Tables 5 and 6).

#### Tremor-frequency band power

Participant- and hemisphere-specific tremor-frequency bands were centred on the individual tremor fundamental frequency (*f₀*) and its second harmonic (2*f₀*). The fundamental frequency was estimated from upper-limb accelerometer recordings acquired during the in-hospital M3 and M6 sessions, with surface EMG used when accelerometry was unavailable or insufficient. Estimates from M3 and M6 were combined to obtain a single *f₀* per participant and limb. Because each thalamic electrode was related to tremor in the contralateral limb, hemisphere-specific *f₀* values were assigned accordingly. The full estimation procedure is described in Supplementary Methods S8, and individual *f₀* values are reported in Supplementary Table 7. Two frequency bands were then defined for each hemisphere: *f₀* ± 1 Hz and 2*f₀* ± 1 Hz. Across the 20 hemispheres, *f₀* ranged from 3.42 to 5.86 Hz (median 4.27 Hz). Power within the fundamental (*f₀*) and second-harmonic (2*f₀*) bands were calculated using the same procedure as for the canonical frequency bands.

#### Aperiodic and periodic spectral parameterisation

For each participant, session, condition and hemisphere, the mean squared amplitude spectrum was parameterised with the FOOOF/specparam^30^ algorithm to separate the aperiodic background from superimposed oscillatory peaks. The model was fitted per channel over 1–40 Hz in fixed aperiodic mode (no knee), with peak-width limits 2–12 Hz, a maximum of three peaks, minimum peak height 0 (no height gate), and a peak threshold of 1.5 standard deviations of the flattened spectrum. Extracted aperiodic features were the exponent and the offset (log₁₀ µVp²), and aperiodic broadband power — the aperiodic fit re-expressed in linear units and summed over the fit-range bins. Aperiodic-corrected band power was calculated as the FOOOF log-space residual (log₁₀(spectrum) − log₁₀(aperiodic fit)) summed over the bins of each band. Periodic peaks were characterised by their centre frequency, amplitude (log₁₀ power above the aperiodic fit) and bandwidth.

### Electrode localisation and impedance assessment

Electrode locations were reconstructed using Lead-DBS,^31^ and the position of the active contact level relative to the ventral lateral posterior nucleus (VLPv), which corresponds to the VIM of Hassler’s nomenclature,^32^ was described from participant-specific HIPS-THOMAS segmentations. Electrode impedance was evaluated at M3 and M6 as an additional technical stability measure, with longitudinal changes assessed using paired non-parametric tests. Full details of electrode localisation and impedance assessment are provided in Supplementary Methods S3 and S9.

### Software

Percept recordings were imported and organised in MATLAB using the Percept toolboxes.^33^ Subsequent analyses were performed in Python, using MNE-Python^34^ for signal processing, FOOOF for spectral parameterisation,^30^ and statsmodels^35^ for linear mixed-effects modelling and multiple-comparison correction. Intraclass correlation coefficients, permutation tests, and bootstrap procedures were implemented in Python following established definitions and procedures. Figures were generated using Matplotlib.^36^ Specific software and package versions are provided in Supplementary Methods S10. Analysis code is available at https://github.com/sahar-allouch/ecolovim-dbs-lfp-home-vs-clinic.

### Statistical analysis

Analyses were performed using bilateral values, calculated for each participant, session and condition as the equal-weight mean of the corresponding left- and right-hemisphere feature values. Canonical and tremor band power, aperiodic broadband power and the aperiodic exponent were natural-log transformed before statistical analysis. Aperiodic offset, aperiodic-corrected band power and beta peak amplitude were retained on their native log₁₀ scale. Beta peak centre frequency and bandwidth were analysed in Hz without transformation.

#### Session-related differences

Systematic differences between M3, home and M6 were assessed using linear mixed-effects models fitted separately for each feature and recording condition (rest and movement, with stimulation OFF and ON). Timepoint was included as a fixed effect and participant as a random intercept, with home as the reference level. This provided direct estimates of M3–home and M6–home differences, while the M3–M6 contrast was derived from the same fitted model. Models were estimated by restricted maximum likelihood.

Inference was based on within-participant permutation testing (5000 permutations), with 95% confidence intervals (CIs) obtained by participant-level bootstrap resampling (2000 resamples). Effects are reported as model coefficients (β), 95% confidence intervals, permutation *P*-values and standardised effect sizes, calculated as *d* = β/√(τ² + σ²), where τ^2^ and σ^2^ denote the between-participant and residual variance components, respectively. For log-transformed features, model coefficients were back-transformed where appropriate to aid interpretation.

Benjamini–Hochberg false discovery rate (FDR) correction was applied separately within each session contrast across all 13 features and four recording conditions, yielding 52 tests per contrast and analysis unit. Statistical significance was defined as FDR-adjusted *P* < 0.05. Models used all available observations for each feature and condition. Full details of the resampling procedures, singular-fit handling, back-transformations and sensitivity criteria are provided in Supplementary Methods S9.

#### Between-session reliability

Between-session agreement was assessed using two-way random-effects, absolute-agreement, single-measure intraclass correlation coefficients (ICCs; ICC(2,1)).^37,38^ ICCs were calculated for M3–home, M3–M6 and home–M6 for each feature and recording condition (rest and movement, with stimulation OFF and ON). Ninety-five per cent confidence intervals were obtained from 5000 participant-level bootstrap resamples. ICCs were classified as poor (<0.50), moderate (0.50–0.75), good (0.75–0.90) or excellent (>0.90).^39^ Complete cases were used for each comparison (*n* = 9 for M3–M6 and *n* = 8–9 for comparisons involving home); beta peak analyses included 7–9 pairs because a peak was required at both sessions.

#### Movement-related modulation

Movement-related modulation was quantified within each participant, session and stimulation state as the paired difference between movement and rest recordings. For log-transformed features, this corresponds to the log ratio of movement to rest.

For each feature, a linear mixed-effects model was fitted to the movement–rest difference, with timepoint (M3, home and M6) and stimulation state (OFF and ON) as fixed effects and participant as a random intercept. Home and stimulation OFF were used as reference levels. The model therefore tested whether the movement effect differed between home and either clinical visit and whether it differed between stimulation OFF and ON. Models were estimated by restricted maximum likelihood.

Movement effects within stimulation OFF and ON were additionally estimated from state-specific models as the average movement–rest contrast across M3, home and M6. These state-specific estimates describe the magnitude of movement modulation within each stimulation state; differences between stimulation states were assessed from the stimulation term in the full model.

Statistical inference was permutation-based, with 95% confidence intervals obtained by participant-level bootstrap resampling. Benjamini–Hochberg false discovery rate correction was applied across the 13 features separately for each model term and for the stimulation-specific movement contrasts. Statistical significance was defined as FDR-adjusted *P* < 0.05. Full details of the permutation and bootstrap procedures are provided in Supplementary Methods S9.

## Results

### Study population

Ten patients with essential tremor treated with bilateral DBS of the VIM/PSA (20 hemispheres) were enrolled (seven women; mean age 63.5 ± 8.1 years, range 46–74; mean disease duration 20.9 ± 12.0 years, range 11–49). Baseline characteristics are given in Supplementary Table 1.

Participant P03 withdrew after M3 and had no subsequent recordings; this participant was therefore excluded from inferential analyses. Participant P04 had no usable home recordings during stimulation OFF and was excluded from clinic–home comparisons under stimulation OFF only; this participant remained included in M3–M6 comparisons and in all analyses under stimulation ON. Consequently, analyses involving home recordings included eight participants under stimulation OFF and nine under stimulation ON.

Across the 18 hemispheres of the nine analysed participants, the active contact level lay on average at *x* = ±14.3 ± 2.3 mm, *y* = −15.2 ± 1.5 mm and *z* = 0.0 ± 2.4 mm in Montreal Neurological Institute (MNI) space (MNI152NLin2009bAsym; right 15.0, −14.7, 1.1; left −13.6, −15.7, −1.0), at the ventral border of the motor thalamus. Electrode reconstructions obtained with Lead-DBS are shown in Supplementary Fig. 1. As the bipolar sensing pair was consistently centred on the stimulating level, the midpoint of the recording contacts corresponded to the same location. On patient-specific HIPS-THOMAS segmentations, both contacts of the bipolar sensing channel lay within the VLPv (≥ 50% of the contact volume) in 6 of the 18 hemispheres, one contact in 5 and neither contact in 7 (Supplementary Methods S3); contacts lying outside the nucleus were mostly located ventral to it, towards the posterior subthalamic area.

Stimulation produced the expected clinical benefit: the total Fahn–Tolosa–Marin score fell from 53.2 ± 11.5 before surgery to 20.5 ± 11.8 at M3 with stimulation ON (Wilcoxon signed-rank test, *P* = 0.002), every patient improving, by 31% to 86% individually (mean 63.1 ± 15.5%). Patients rated the home procedure as easy to carry out, 8.7 ± 1.1 on a 10-point visual analogue scale (range 7–10).

The analysed dataset comprised in-clinic recordings at M3 and M6, each condition yielding one spectrum per hemisphere from on average 61–105 s of usable signal (Supplementary Table 4), and 770 home event snapshots of 30 s (≈6.4 h of recording; 179–203 per condition; median 87 per participant; Supplementary Table 3), which were averaged within participant and condition before analysis. Fig. 2 provides an overview of the resulting spectral profiles. Group-level spectra at M3 and M6 were closely overlapping across all four conditions, whereas home spectra were comparable to clinic spectra at lower frequencies but showed progressively lower power at higher frequencies, consistent with a steeper aperiodic slope (Fig. 2A). At the participant level, oscillatory features (alpha, beta and tremor-frequency power) showed relatively stable trajectories across sessions, whereas the aperiodic exponent and offset increased at home before returning towards M3 values at M6 (Fig. 2B; remaining features in Supplementary Figs 2 and 3; descriptive statistics in Supplementary Table 8). These differences are examined quantitatively in the following sections.

### Oscillatory features were largely preserved between home and clinic

No oscillatory feature differed systematically between home and clinic recordings after correction for multiple comparisons (Fig. 3 and Supplementary Fig. 5). This held for canonical band power, tremor power, and periodic beta features.

For canonical band power, clinic–home differences were small and of the same order as those between the two supervised clinical visits (median |*d*|: 0.30 for M3–home, 0.37 for M6–home, and 0.22 for M3–M6). Beta power was higher in clinic in all eight clinic–home comparisons (median *d* = 0.58), but this did not survive correction for multiple comparisons (minimum *P_FDR_* = 0.068).

**Figure 2.**
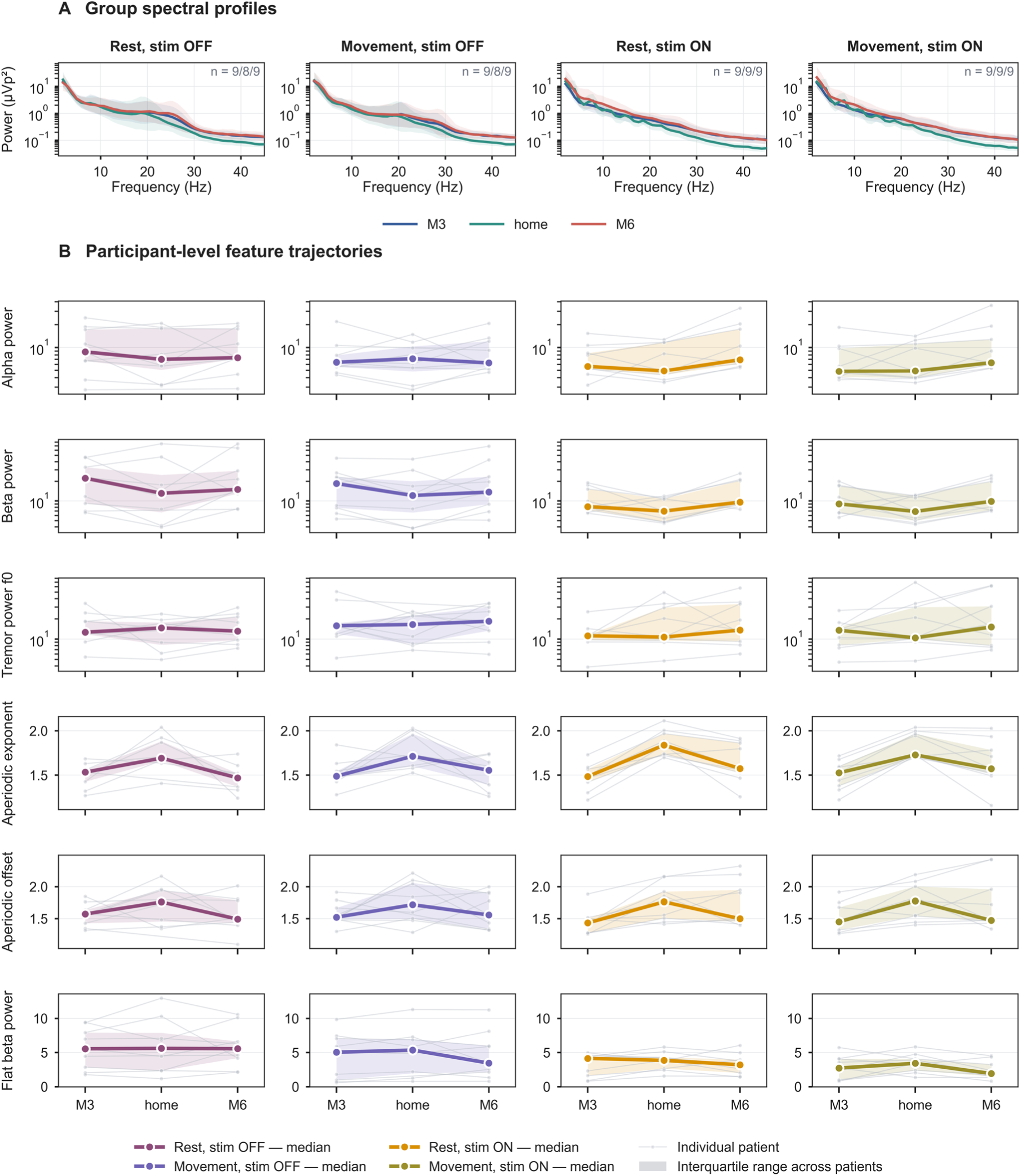
Group spectral profiles and participant-level trajectories across sessions. **(A)** Bilateral LFP power spectra at M3, home and M6 during rest and movement with stimulation OFF and ON. Lines show group spectral profiles and shading shows between-participant variability. **(B)** Participant-level trajectories across M3, home and M6 for selected features: alpha, beta and tremor-frequency power, aperiodic exponent, aperiodic offset and aperiodic-corrected beta power. Other features trajectories can be seen in the Supplementary Results (Supplementary Figs 2 and 3). Coloured lines and points show medians and shaded regions show interquartile ranges; grey lines show individual participants. LFP = local field potential.

Tremor power was similarly stable, with clinic–home differences no larger than those between the two clinical visits (median |*d*|: 0.28 for M3–home and 0.30 for M6–home vs. 0.37 for M3– M6; minimum clinic–home *P* = 0.089).

Periodic beta features, including peak centre frequency, bandwidth, peak power, and aperiodic-corrected power, also showed no clinic–home difference that survived correction (median |*d*|: 0.29 for M3–home and 0.25 for M6–home; minimum *P_FDR_* = 0.16), although these differences were larger than those between the two clinical visits (median |*d*| = 0.09 for M3–M6). Beta peaks were detected in 89–100% of usable recordings in each hemisphere across sessions.

Differences were generally larger with stimulation ON than OFF (median |*d*| = 0.42 for stimulation ON vs. 0.19 for stimulation OFF across all oscillatory features and session pairs), including between the two clinical visits (median |*d*| = 0.56 for stimulation ON vs. 0.09 for stimulation OFF).

### Home recordings showed a selective shift in aperiodic spectral structure

The aperiodic exponent was the only feature to show systematic home–clinic differences after correction for multiple comparisons, with four clinic–home contrasts surviving correction (Fig. 3 and Supplementary Fig. 5); 18 of 104 clinic–home contrasts were nominally significant.

The aperiodic exponent was consistently higher at home, indicating a steeper 1/f spectral slope. The direction of this effect was consistent across rest and movement and with stimulation both OFF and ON, occurring in all eight clinic–home comparisons (all uncorrected *P* ≤ 0.008; *P_FDR_* ≤ 0.068; Fig. 2A and Supplementary Fig. 3). Effect sizes were large relative to both M3 (median *d* = −1.55) and M6 (median *d* = −1.24) and exceeded the difference between the two clinical visits (median |*d*| = 0.41). The offset showed the same directional tendency, being higher at home in all eight comparisons, although the effect was more pronounced relative to M3 (median *d* = −0.71) than M6 (median *d* = −0.29) and did not survive correction (minimum *P_FDR_* = 0.055).

By contrast, aperiodic broadband power showed no consistent home–clinic difference. The M6–home difference (median *d* = 0.41) was comparable in magnitude to the change between M3 and M6 (median |*d*| = 0.47), consistent with session-related variability rather than a systematic home–clinic shift.

Electrode impedance remained stable between M3 and M6, with no hemisphere- or contact-level difference surviving correction for multiple comparisons. Detailed results are presented in Supplementary Results S11 and Supplementary Fig. 6.

**Figure 3.**
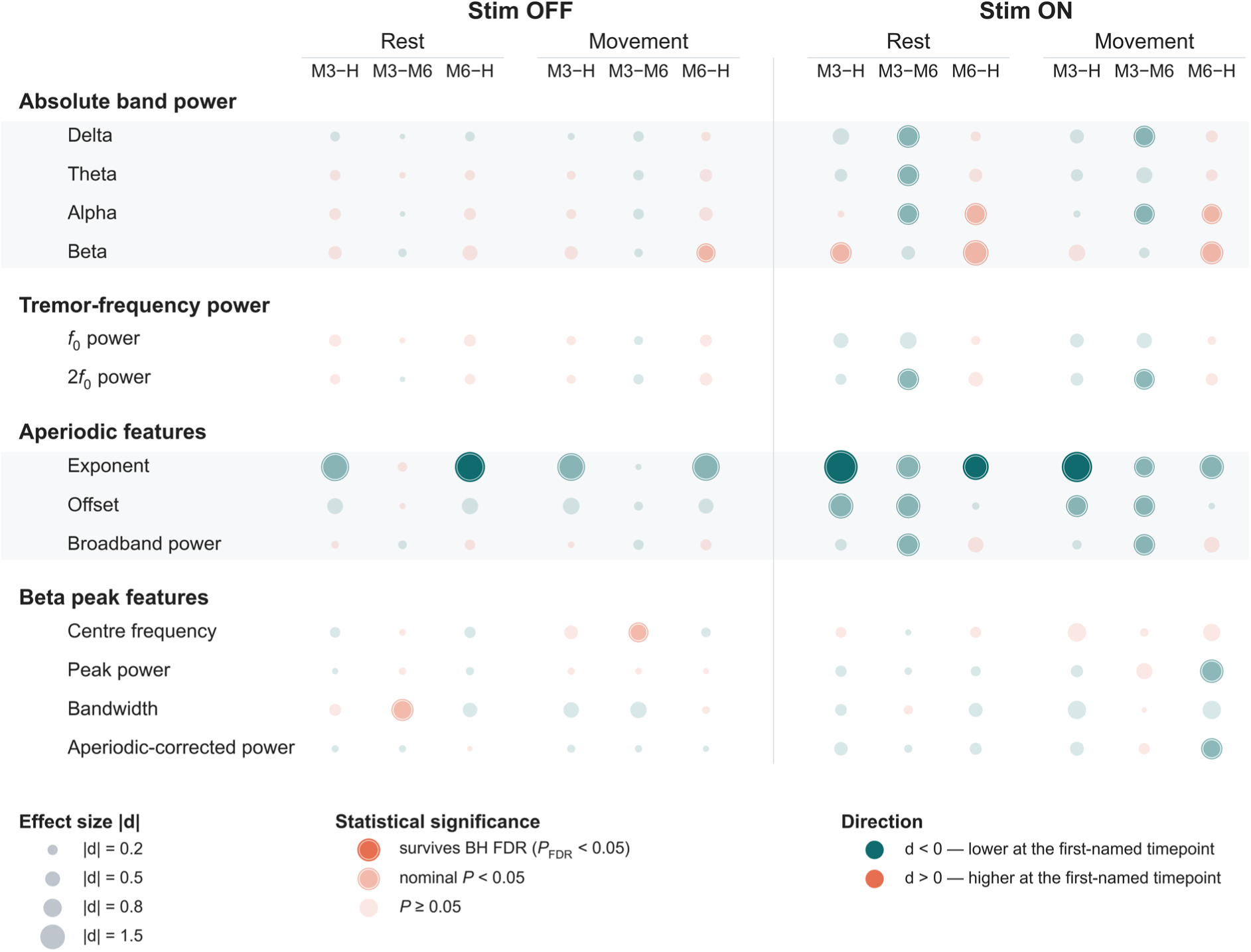
Standardised differences between recording sessions in bilateral LFP features. Cohen’s *d* values are shown for M3–home, M3–M6 and M6–home contrasts during rest and movement with stimulation OFF and ON. Symbol size represents effect magnitude (|*d*|). Teal indicates negative effects (*d* < 0; lower values at the first-named session), and orange indicates positive effects (*d* > 0; higher values at the first-named session). Symbol opacity denotes statistical significance: dark symbols indicate effects surviving Benjamini–Hochberg false-discovery-rate correction (*PFDR* < 0.05), intermediate symbols nominal significance (*P* < 0.05), and pale symbols *P* ≥ 0.05. H = home.

### Between-session reliability varied across features and stimulation states

Between-session reliability varied substantially across feature types and stimulation states (Fig. 4 and Supplementary Fig. 4). Across all features, median ICC(2,1) was 0.47 for M3– home, 0.55 for M6–home and 0.62 for M3–M6.

Periodic beta features showed the highest reliability, particularly during stimulation OFF. Aperiodic-corrected beta power, peak power and peak centre frequency showed moderate-to-excellent clinic–home agreement during stimulation OFF (ICC ranges 0.66–0.96, 0.58–0.91 and 0.63–0.90, respectively), with generally lower agreement during stimulation ON (0.46– 0.75, 0.03–0.61 and 0.02–0.79, respectively). Peak bandwidth was less reliable in both states, with clinic–home ICCs of 0.17–0.62 during stimulation OFF and −0.40 to 0.34 during stimulation ON.

**Figure 4.**
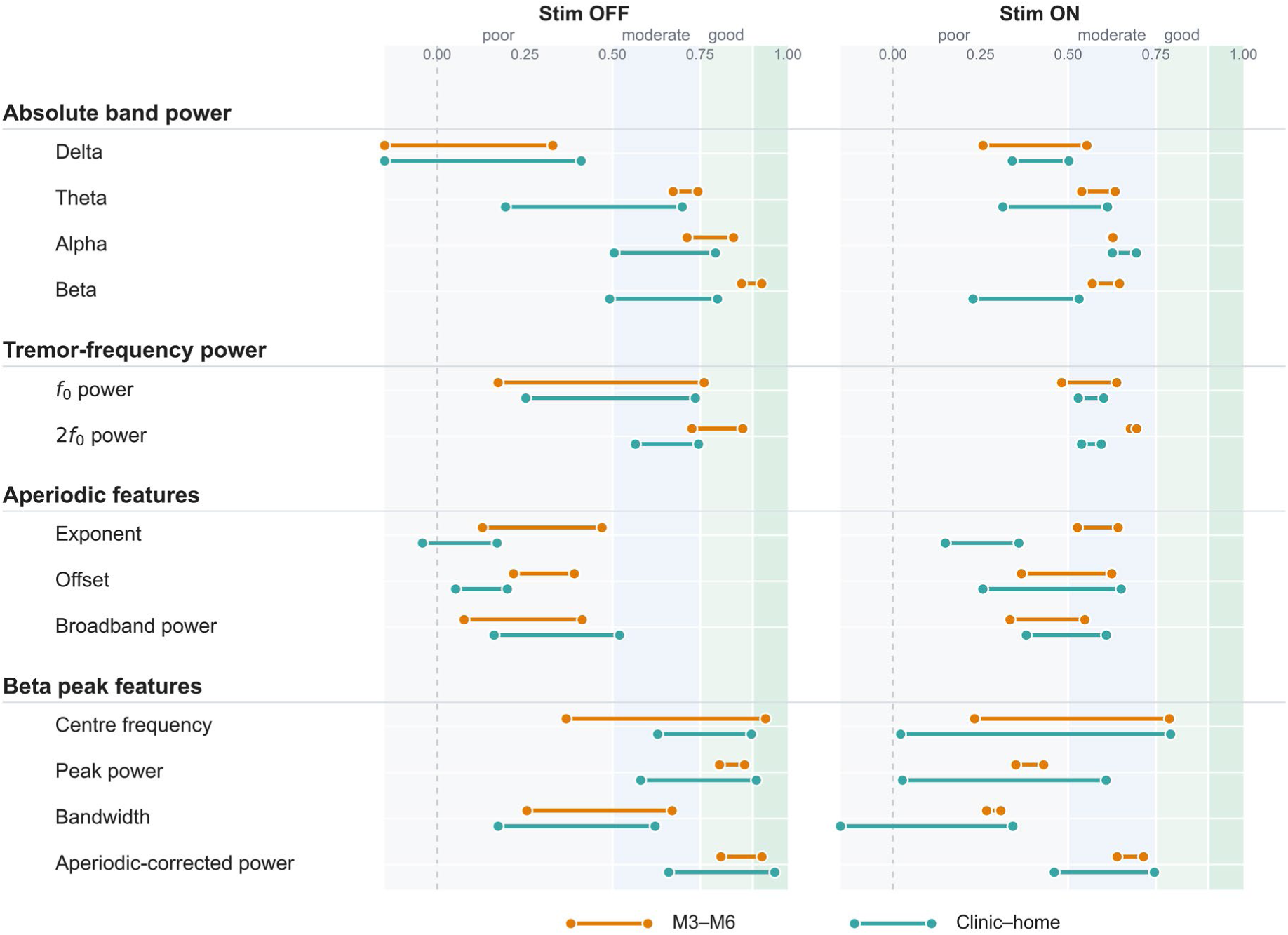
Between-session reliability of bilateral LFP features. ICC(2,1) ranges across behavioural conditions are shown separately for stimulation OFF and ON. Orange ranges represent clinic–clinic reliability between M3 and M6, and teal ranges represent clinic–home reliability (M3–home and M6–home). Background shading denotes conventional ICC reliability categories. ICC = intraclass correlation coefficient.

Tremor-frequency power showed moderate clinic–home agreement overall (median ICCs 0.56 for M3–home and 0.58 for M6–home, versus 0.69 for M3–M6). Among canonical bands, alpha and beta were the most reliable during stimulation OFF (clinic–home ICCs 0.50–0.79 and 0.49–0.80, respectively), whereas theta was more variable (0.19–0.70) and delta showed the lowest agreement (−0.26 to 0.41). During stimulation ON, alpha remained moderately reliable (0.63–0.69), whereas beta agreement decreased (0.23–0.53); theta and delta ranged from 0.31–0.61 and 0.34–0.50, respectively.

Aperiodic features showed low between-session reliability, particularly during stimulation OFF. Median clinic–home ICCs were 0.04 for the exponent, 0.12 for the offset and 0.19 for broadband power, during stimulation OFF, with similarly limited reliability between the two clinical visits. Reliability generally improved during stimulation ON, particularly for offset and broadband power. The exponent remained less reliable between clinic and home than between the two clinical visits; however, ICC(3,1) exceeded ICC(2,1) for clinic–home comparisons (0.50–0.70 versus 0.15–0.36; stimulation ON), indicating better preservation of between-participant ranking than absolute agreement in the presence of the systematic home–clinic shift.

### Movement modulated oscillatory beta activity during stimulation OFF

With stimulation OFF, movement produced a consistent reduction in beta power across all three sessions (Fig. 5A), extending approximately from 13 to 25 Hz. Beta power decreased by 18% (β = −0.20 ln, 95% CI [−0.29, −0.10], *P* = 0.004, *P_FDR_* = 0.025), with no evidence that this effect differed across sessions (movement × session, all *P_FDR_* ≥ 0.89; Fig. 5B and Supplementary Table 9). Power at the tremor frequency tended to increase with movement (β = 0.18 ln, [0.06, 0.38], *P* = 0.020), although this did not survive correction (*P_FDR_* = 0.064). Delta, theta, alpha and second-harmonic tremor power were not significantly modulated (all |β| ≤ 0.14 ln, all *P* ≥ 0.09, all *P_FDR_* ≥ 0.19).

The absolute beta power reduction reflected changes in periodic rather than aperiodic features. Aperiodic exponent, offset and broadband power remained unchanged (all changes ≤ 10%, all *P_FDR_* ≥ 0.28), whereas aperiodic-corrected beta power decreased markedly (β = −1.15 log₁₀ units, [−1.76, −0.60], *P* = 0.004, *P_FDR_* = 0.025). Movement also shifted the beta peak towards higher frequencies (β = 1.10 Hz, [0.56, 1.64], *P* = 0.008, *P_FDR_* = 0.034). Beta peak amplitude showed a smaller reduction that did not survive correction (β = −0.09 log₁₀ units, [−0.14, −0.03], *P* = 0.031, *P_FDR_* = 0.081).

Movement-related effects were consistent between home and clinic recordings. None of the 13 features showed a setting-related difference in movement response after correction for multiple comparisons (all *P_FDR_* ≥ 0.45).

### Stimulation attenuated movement-related beta modulation

Stimulation markedly attenuated movement-related beta suppression, with movement–rest difference spectra becoming largely flat across the beta range during stimulation ON (Fig. 5A). Absolute beta power no longer changed with movement (β = −0.00 ln, 95% CI [−0.06, 0.06], *P* = 0.94; Fig. 5B and Supplementary Table 10), and the movement effect differed significantly between stimulation states (β = 0.20 ln, [0.12, 0.27], *P* = 0.0002, *Pfdr* = 0.003).

Periodic beta activity showed a similar attenuation. The movement-related decrease in aperiodic-corrected beta power was 0.65 log₁₀ units smaller during stimulation ON than OFF ([0.22, 1.07], *P* = 0.004, *P_FDR_* = 0.027). A residual decrease remained during stimulation ON (β = −0.49 log₁₀ units, [−0.83, −0.11], *P* = 0.047) but did not survive correction (*P_FDR_* = 0.30). No other spectral or aperiodic feature showed a movement-related change during stimulation that survived correction (all *P_FDR_* ≥ 0.30).

**Figure 5.**
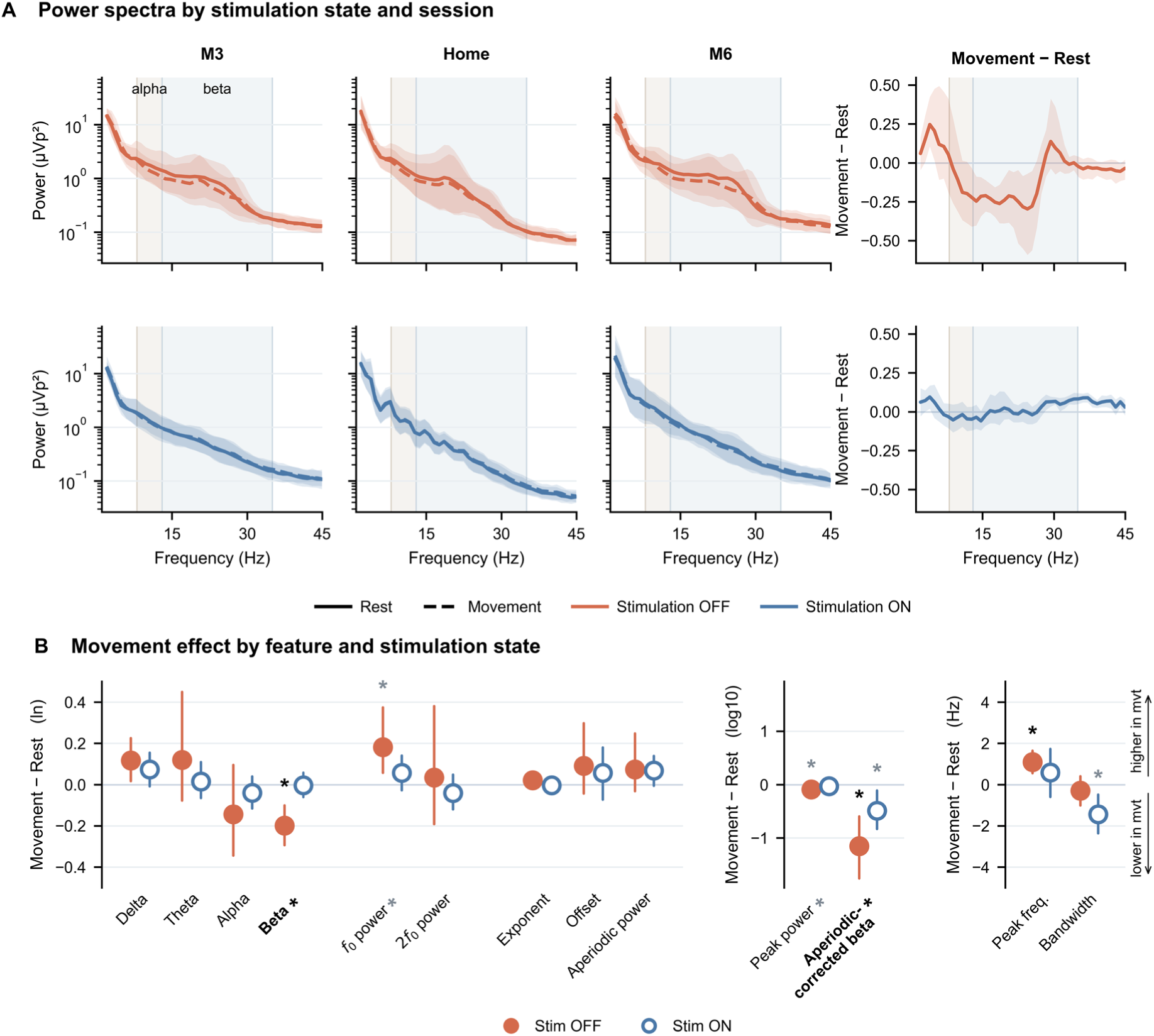
Movement-related modulation of thalamic LFP activity. **(A)** Bilateral power spectra during rest and movement at M3, home and M6, shown separately for stimulation OFF and ON, with corresponding movement– rest difference spectra. **(B)** Estimated movement–rest effects for spectral features during stimulation OFF and ON; points and error bars show model estimates and 95% confidence intervals. Stars above estimates denote within-stimulation-state movement effects, whereas stars beside feature labels denote movement × stimulation interactions. Black stars indicate effects surviving false-discovery-rate correction and grey stars nominal significance.

### Beta features combined reliability and movement sensitivity

Fig. 6 summarises these two properties by plotting, for each of the 13 features, clinic–home reliability against the movement–rest effect. With stimulation OFF, only beta power, aperiodic-corrected beta power and beta peak centre frequency combined at least moderate reliability with a movement effect that survived correction. With stimulation ON, no feature met both criteria.

**Figure 6.**
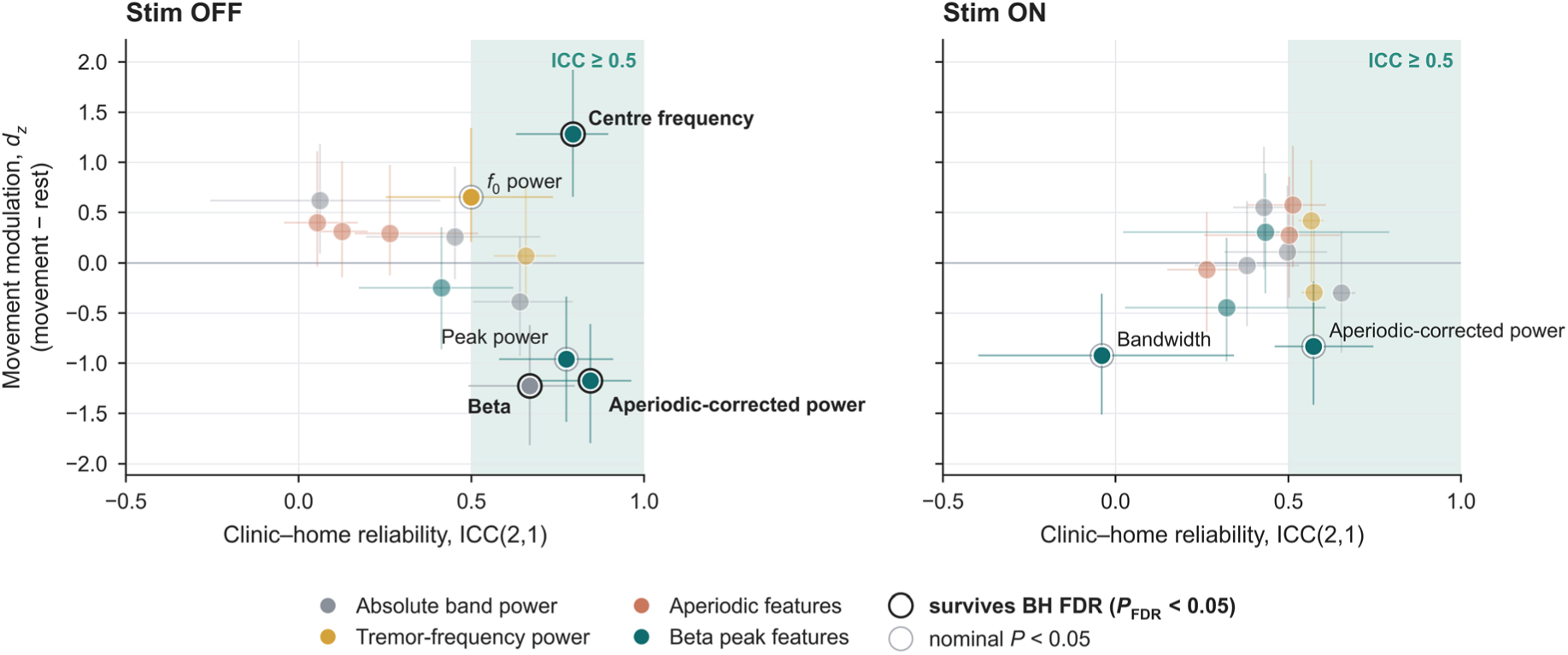
Identifying promising LFP biomarkers in essential tremor. Clinic–home reliability (ICC(2,1)) is plotted against standardised movement–rest effects (*dz*) for 13 spectral features with stimulation OFF and ON. Colours distinguish feature categories. Horizontal bars show ICC ranges across recording comparisons, and vertical bars show 95% confidence intervals for movement effects. Black outlines indicate effects surviving false-discovery-rate correction (*PFDR* < 0.05), and grey outlines indicate nominal significance (*P* < 0.05). Background shading indicates ICC ≥ 0.5. Periodic beta features showed the most favourable combination of clinic–home reliability and movement sensitivity, particularly with stimulation OFF, highlighting their potential as candidate biomarkers for adaptive DBS. DBS = deep brain stimulation; ICC = intraclass correlation coefficient; LFP = local field potential.

## Discussion

In this study, we investigated whether thalamic spectral features proposed as biomarkers for aDBS in essential tremor when acquired at home, are comparable to in-clinic measurements, and whether they exhibit similar movement-related modulation across recording settings. To this end, we first examined the feasibility of unsupervised home thalamic recordings in patients with essential tremor. We then characterised the agreement between spectral features derived from home and in-clinic recordings using two complementary approaches: linear mixed-effects models to identify systematic between-session differences, and intraclass correlation coefficients to assess the reliability of individual feature values across sessions. Finally, we used linear mixed-effects models of the movement–rest difference to test whether movement-related modulation varied across recording settings and stimulation states.

Patients completed weekly sessions and rated the procedure as easy to perform. This is an important step towards clinical translation, since candidate aDBS biomarkers must ultimately be characterised beyond supervised clinical setting. In Parkinson’s disease, subthalamic and pallidal signals acquired in daily life are increasingly used to inform adaptive stimulation strategies,^25,40^ and recent work has extended this approach to gait-responsive stimulation during everyday activities.^41^ Comparable evidence remains scarce in essential tremor, where thalamic activity has mostly been studied in small cohorts under supervised conditions. Our study helps to fill this gap by assessing the reliability and movement-related modulation of candidate thalamic biomarkers during patient-managed home recordings.

Several key findings emerged. Oscillatory features were broadly consistent between home and clinic, whereas the aperiodic background showed a systematic shift towards steeper spectra at home. Reliability differed markedly across features: periodic beta measures showed the highest clinic–home reliability, approaching that observed between clinic visits, whereas aperiodic measures were less reliable. Stimulation was associated with lower beta reliability, highlighting the dependence of these measures on stimulation state. Movement-related beta suppression was observed across recording settings and was predominantly associated with periodic activity rather than changes in the aperiodic background. However, this modulation was markedly attenuated during stimulation.

Across oscillatory features, home–clinic differences were generally comparable in magnitude to those between the two clinic visits. Absolute beta power was a notable exception, being higher in the clinic in all eight contrasts, although none survived correction for multiple comparisons. In contrast, aperiodic-corrected beta power, which measures beta activity relative to the fitted aperiodic background, tended to be higher at home (seven of eight contrasts). These opposing trends may reflect the systematic shift in aperiodic activity between settings, as a flatter spectrum can increase absolute beta power without a corresponding increase in periodic beta activity.^42^ This distinction has implications for aDBS: although oscillatory features may be reproducible across settings, current device controllers measure total band power, including its aperiodic contribution. Consequently, beta-power thresholds calibrated in the clinic may not transfer directly to home recordings.

The aperiodic component, the broadband 1/f activity on which oscillatory peaks sit, is characterised by its exponent and offset.^30^ The exponent and offset were consistently higher at home across all eight clinic–home comparisons, with large differences relative to both M3 and M6 that exceeded those between clinic visits. On the other hand, aperiodic broadband power showed no significant difference between settings. Together, these findings suggest a change in spectral slope rather than a uniform shift in broadband power, with relatively greater low-frequency and lower high-frequency aperiodic activity at home. A similar dissociation has been reported in the subthalamic nucleus following levodopa administration, where the exponent increased without a corresponding change in aperiodic broadband power.^42^ A postoperative trajectory is unlikely to fully explain this pattern. Because home recordings were acquired between M3 and M6, a monotonic postoperative change would be expected to produce intermediate values rather than consistently higher values at home than at either clinic visit. Previous subthalamic recordings have also shown no significant changes in exponent or offset between one to three months and one year after implantation.^43^ Nevertheless, changes in aperiodic exponent can arise from multiple mechanisms,^42,44^ and the present finding remains descriptive and should not be interpreted as evidence of altered excitation–inhibition balance.

Three explanations remain possible. The first is recent stimulation history: home stimulation-OFF recordings followed an overnight interruption, whereas clinic recordings were obtained after only a 5-min washout. In the subthalamic nucleus, stimulation increases both the aperiodic exponent^45,46^ and offset.^46^ Any residual stimulation effect after the shorter clinic washout would therefore be expected to increase these parameters in the clinic rather than at home, contrary to the observed pattern. The second possibility is vigilance. Home stimulation-OFF recordings were obtained on waking, and it has been reported that both the aperiodic exponent and offset increase during the transition from wakefulness to sleep in cortical and subthalamic recordings.^47^ Then, we hypothesise that residual drowsiness after waking could therefore account for the steeper spectra and higher offset observed at home, although this mechanism has not yet been established in the thalamus. Lastly, although clinic spectra were processed to approximate the device output, differences between offline Welch estimates and on-chip FFT snapshots could affect the fitted aperiodic parameters. Whether these differences contributed to the systematic home–clinic shift remains uncertain.

Between-session reliability differed markedly across features. Periodic beta measures were the most reproducible, with clinic–home ICCs approaching those observed between clinic visits, whereas the aperiodic exponent and offset were poorly reliable, a pattern also reported in subthalamic recordings in Parkinson’s disease.^27^ Aperiodic broadband power, proposed as a within-patient marker of subthalamic activity,^42^ was poorly reliable with stimulation OFF and only moderately reliable with stimulation ON. Stimulation, in turn, reduced the reliability of all five beta features, both between clinic and home and between the two clinic visits, indicating that this decline reflected the stimulated state rather than the home setting.

These findings warrant two caveats. First, ECG contamination may have contributed to the lower beta reliability during stimulation, when the device case serves as the reference; right-sided implantation, as in our cohort, reduces but does not abolish this risk.^48^ ECG contamination was more apparent in our clinic recordings during stimulation ON but could not be assessed at home, where time series were unavailable, and a subthalamic study found no loss of beta-peak reliability during stimulation once the recording configuration was matched and ECG contamination removed.^27^ Second, low ICCs should not be equated with random measurement noise. ICCs depend on between-participant variability as much as on measurement error,^39^ and absolute-agreement estimates are further penalised by systematic shifts. With stimulation, the between-participant standard deviation of beta-amplitude measures fell to 25–51% of its OFF value, consistent with the suppression of subthalamic beta power by stimulation,^49^ which mechanically lowers ICCs. For the aperiodic exponent, between-participant ranking was largely preserved despite poor absolute agreement (ICC(3,1) 0.50–0.70 versus ICC(2,1) 0.15–0.36 with stimulation ON), pointing to a consistent setting-related bias that could, in principle, be corrected by calibration.

Movement-related thalamic beta suppression, a promising candidate signal for aDBS, was observed across recording settings. With stimulation OFF, beta power decreased during the postural task, with no significant differences in the beta response between home and either clinic visit. Averaged across sessions, movement reduced absolute beta power (β = −0.20 ln, *P_FDR_* = 0.025) and corrected beta power (β = −1.15 log₁₀, *P_FDR_* = 0.025), while increasing beta peak frequency (+1.10 Hz, *P_FDR_* = 0.034). No corresponding modulation was detected in aperiodic parameters (all *P_FDR_* ≥ 0.28), suggesting that beta suppression predominantly reflected changes in periodic activity rather than the aperiodic background. These findings are consistent with previous reports of thalamic alpha and low-beta suppression during cued button presses in externalised recordings,^50^ and sustained beta modulation during posture and action over 24 months of chronic recording.^19^ By contrast, tremor-frequency power showed no significant modulation during the postural task (all *P_FDR_* ≥ 0.06). This dissociation suggests that beta activity primarily reflects motor state rather than tremor expression, consistent with thalamic decoding studies in which low-beta activity tracked voluntary movement whereas theta activity tracked postural tremor.^20^ The absence of robust tremor-frequency modulation may reflect the spatial variability of thalamic tremor-related activity, as previous work has shown that its relationship with tremor depends on electrode location and the distribution of tremor-related neurons.^16^

Stimulation markedly attenuated movement-related beta modulation. Absolute beta power no longer decreased during the postural task, with a significant reduction in the movement response relative to stimulation OFF (*P_FDR_* = 0.003). Although a smaller movement-related decrease persisted in corrected beta power (−0.49 log₁₀ (95% CI −0.83 to −0.11)), this residual modulation does not establish reliable movement detection during stimulation. This distinction is particularly relevant for movement-triggered aDBS, in which movement onset and the return to rest are detected under different stimulation states. In a previously tested thalamic aDBS system for essential tremor, stimulation was activated upon movement detection and discontinued when movement ceased.^11^ Movement onset was therefore detected during stimulation OFF, when beta modulation was strongest in our recordings, whereas the return to rest had to be detected during stimulation ON, when beta modulation was substantially attenuated. This challenge has been recognised in earlier systems: stimulation-related interference prevented reliable detection of movement cessation with a previous-generation device,^12^ while He et al. used separate movement decoders for stimulation OFF and ON.^11^ Similar stimulation-dependent changes in beta activity have been shown to impair motor-state classification in Parkinson’s disease.^42^ These findings highlight the need to evaluate movement onset and cessation separately, with detection thresholds established under their respective stimulation states.

The stability of the monitored frequency band presents a further challenge for thalamic aDBS. Current implantable controllers typically monitor power within a narrow frequency band defined during calibration, often centred on an individual beta peak in Parkinson’s disease.^51^ In our recordings, beta peak frequency increased during the postural task, the magnitude of this shift varied across sessions, and peak bandwidth showed poor reliability. Such variability may compromise fixed-band monitoring, even when movement-related beta modulation remains consistent. Similarly, a chronic closed-loop study in essential tremor identified shifts in the centre frequency and bandwidth of thalamic oscillations as major obstacles to reliable control, requiring frequent recalibration.^22^ Integrating beta power over a wider frequency range or dynamically adjusting the monitored band to track the beta peak may reduce sensitivity to these shifts, although neither approach was evaluated here. Complementary signals may also improve the robustness of adaptive control. Sensorimotor cortical activity has shown greater long-term stability than thalamic signals^22^ and has supported closed-loop stimulation with reduced energy consumption.^12^ Peripheral sensors have also enabled responsive stimulation with tremor control comparable to continuous stimulation,^52^ supporting the potential of multimodal approaches that combine neural and peripheral signals.^53^

### Limitations

The sample of nine participants limits precision and precludes formal equivalence testing. Home spectra were averaged across weekly recordings acquired between M3 and M6 and compared with single clinic sessions; therefore, the reported agreement reflects the mean home signal rather than week-to-week repeatability or the reliability of a single snapshot. Home task performance was patient-reported and without objective kinematic verification. Wearable sensors, which capture aspects of tremor burden not reflected by clinical scales,^54^ could provide such information in future studies. Tremor severity was assessed only at M3, limiting inference on associations between spectral features and longitudinal changes in tremor clinical severity. Stimulation history and recording conditions differed between settings by design, and minor stimulation adjustments were required in some participants between visits. A residual stun effect, reported to persist up to three months after thalamic surgery in a randomised trial,^6^ may therefore have influenced M3 recordings and was not independently tracked. However, the stability of measure between M3 and M6 rules out this hypothesis. ECG contamination could not be quantified in the home recordings. Finally, because the sensing pair was determined by the active stimulation contact, recording locations varied across hemispheres: both contacts of the sensing channel lay within the VLPv, used here as an imaging surrogate of the VIM, in only 6 of the 18 analysed hemispheres, at least one contact lay within it in 11. The stability of electrode impedances argues against a drift in recording quality as the primary explanation for session-related effects.

This study demonstrates the feasibility of patient-managed thalamic LFP recordings in essential tremor and provides a detailed characterisation of thalamic spectral features across recording settings and stimulation states. Periodic beta measures showed relatively high clinic– home reliability and consistent movement-related modulation; stimulation, however, attenuated both, exposing a key challenge for adaptive control. Thalamic beta thus remains a meaningful marker of motor state in daily life, but not yet a stand-alone control signal during stimulation; complementary cortical or peripheral inputs, together with validation in the stimulated state, will be needed before thalamic adaptive stimulation can be broadly implemented in essential tremor.

## Data availability

All code used for data processing and analysis is openly available at https://github.com/sahar-allouch/ecolovim-dbs-lfp-home-vs-clinic. The datasets generated and analysed in this study will be made openly available as a separate data publication. Prior to that release, they are available from the corresponding author upon reasonable request.

## Acknowledgements

We thank the patients and their families for their participation in this study.

We also thank Gaetano Leogrande (Medtronic) for his technical expertise and support regarding on-device computation of the Percept neurostimulator.

## Supplementary material

Supplementary material is available online.

