## Supplemental for "Ecological characterisation of frequency-domain local field potential biomarkers in essential tremor"

### Supplementary material

#### 1. Supplementary Methods

##### S1. Participants

We prospectively studied ten patients with severe, disabling, medication-refractory essential tremor treated with deep brain stimulation (DBS) of the ventral intermediate nucleus (VIM) of the thalamus or the posterior subthalamic area (PSA) as part of standard clinical care. Inclusion criteria were: a diagnosis of essential tremor fulfilling the 2018 consensus criteria of the International Parkinson and Movement Disorder Society[1]; treatment with the Percept PC/RC system (Medtronic, Minneapolis, MN); age 18–75 years; a normal brain MRI; preserved global cognition (Mattis Dementia Rating Scale ≥130 and/or Montreal Cognitive Assessment ≥24); affiliation to the national health-insurance system; and written informed consent. Exclusion criteria were major depressive disorder (Beck Depression Inventory >20); marked cerebral atrophy or extensive MRI signal abnormality; pregnancy or breastfeeding; inability to give informed consent; and legal guardianship. Patients continued their usual anti-tremor medication during the study. The study was approved by the Comité de Protection des Personnes Nord Ouest IV (Lille, France; approval no. 22.04558.000153), sponsored by Bordeaux University Hospital, and supported by institutional funding (CHU de Bordeaux, AOI 2022). All participants gave written informed consent. Trial registration: NCT05671848.

Supplementary Table 1. Participant demographic and clinical characteristics. Age range, sex, disease duration, Fahn Tolosa and Marin[2] (FTM) scores before surgery and at M3 (ON-DBS), and FTM-defined symptom-dominant side are shown. Dashes indicate tied lateralized FTM subscores (Left-Right).

| | | | | FTM, total score | | Symptom dominant side[b] | |
|---|---|---|---|---|---|---|---|
| **ID** | **Age range, years** | **Sex** | **DD[a], years** | **Preop** | **M3** | **Preop** | **M3** |
| P01 | 66–70 | M | 32 | 49 | 16 | — | R |
| P02 | 46–50 | F | 21 | 36 | 5 | L | R |
| P03 | 61–65 | F | 17 | 51 | 22 | L | L |
| P04 | 56–60 | F | 48 | 70 | 48 | R | R |
| P05 | 56–60 | F | 11 | 58 | 32 | L | L |
| P06 | 56–60 | F | 29 | 63 | 20 | L | — |
| P07 | 61–65 | M | 11 | 41 | 15 | R | — |
| P08 | 71–75 | F | 11 | 63 | 13 | R | L |
| P09 | 66–70 | F | 12 | 61 | 17 | R | — |
| P10 | 66–70 | M | 10 | 38 | 17 | L | — |

[a] DD, disease duration.
[b] Worse side by FTM subscore at the timepoint headed (preop or M3 stim-ON); a dash marks a tied subscore.

#### S2. Surgical procedure and device

Electrodes were implanted under general anaesthesia using a previously published targeting method[3]. A Leksell stereotactic frame (Elekta, Stockholm, Sweden) was positioned, and stereotactic targeting used a 3-Tesla Canon MRI scanner (Canon Medical Systems, Otawara, Japan) acquired preoperatively. Two directional leads (B33005, Medtronic) were implanted in the VIM under intraoperative imaging guidance (O-arm, Medtronic), and a bidirectional implantable pulse generator (Percept PC/RC, Medtronic) was placed in the right subclavicular or abdominal region during the same procedure.

#### S3. Structural imaging and electrode localization

All patients underwent preoperative brain MRI as part of standard care, including a high-resolution 3D T1-weighted sequence and a white-matter–nulled sequence used for thalamic segmentation and anatomical targeting. A postoperative cranial CT was acquired 48–72 h after surgery to verify lead placement. Electrodes were localized with Lead-DBS v3.2 (lead-dbs.org)[4,5]. In brief, postoperative CT was linearly coregistered to the preoperative MRI with Advanced Normalization Tools (ANTs)[6]; coregistrations were visually inspected and refined where necessary, and a brain-shift correction was applied. Preoperative volumes were normalized into ICBM 2009b NLIN asymmetric ("MNI") space using multispectral normalization, and warp fields were manually refined with WarpDrive[7]. Thalamic nuclei were defined using the HIPS-THOMAS atlas[8,9]. DBS contacts were reconstructed manually. Group-level visualizations used the Lead Group toolbox[10]. All coregistrations and normalizations were visually inspected for quality control and refined manually when needed. For each contact level, the proportion of contact volume located within the ventral part of the ventral lateral posterior nucleus (VLPv) was quantified. In the Hirai and Jones nomenclature adopted by the Morel atlas and THOMAS, VLPv is the cerebellar-recipient territory that corresponds approximately to Hassler's VIM, and it is commonly used as its anatomical surrogate in imaging studies[11–13]. To describe electrode placement, the overlap between each contact and the VLPv was computed in each participant's native space, on the brain-shift-corrected reconstruction, as the proportion of the contact volume lying within the participant's own VLPv segmentation. Each bipolar sensing channel was characterised by the VLPv overlap of its two contacts; lead orientation was not required, as directional levels were always sensed as a whole ring in our recordings. Using a 50% threshold, both contacts lay within the VLPv in 6 of the 18 analysed hemispheres, one contact in 5 and neither in 7.

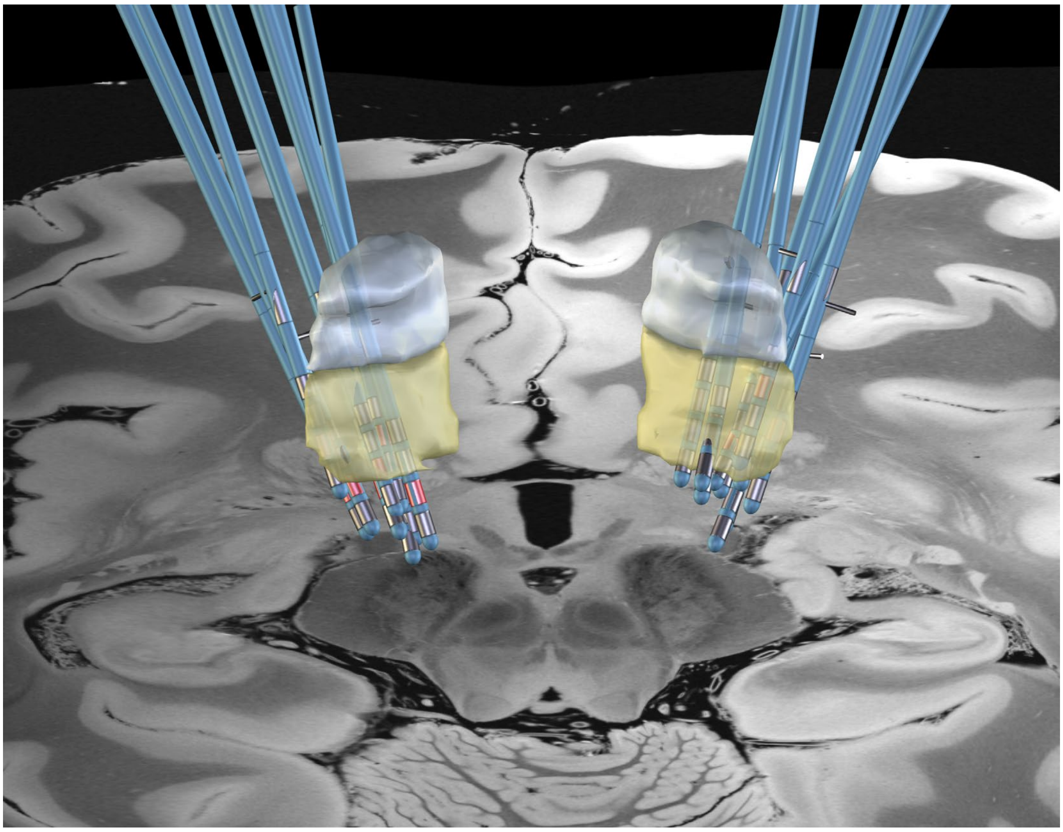

Supplementary Fig. 1. Reconstruction of bilateral DBS electrodes relative to the ventral lateral posterior thalamic nucleus. Electrode leads (blue) are shown with the ventral (VLPv, corresponding to Hassler's Vim; yellow) and dorsal (VLPd; grey) subdivisions of the nucleus from the THOMAS atlas[8,9], displayed on the 7 T ex vivo 100 µm template[14] in MNI space. Active contacts are highlighted in red.

#### S4. Clinical assessment

Tremor severity was rated with the Fahn–Tolosa–Marin scale preoperatively as part of the routine clinical assessment and at the inclusion visit three months after surgery (M3). The scale comprises 21 clinician-rated items scored from 0 to 4, covering tremor severity by body segment and condition (rest, posture, intention), tremor during specific tasks (writing, drawing, pouring, speaking), and the impact of tremor on activities of daily living; higher scores indicate more severe tremor.

#### S5. Stimulation parameters

Stimulation settings were adjusted as clinically indicated during follow-up and systematically recorded at each visit. The settings in place during the M3 and M6 recordings were read from the Percept session reports and are listed per patient and hemisphere in Supplementary Table 2, where intensity is given as the current per active contact. All configurations were monopolar. At M3 (10 patients, 20 hemispheres), the total current delivered per hemisphere, i.e. the sum over active contacts, was 1.7 ± 0.6 mA (range 0.9–3.6), the pulse width 59 ± 21 µs (40–120) and the frequency 140 ± 24 Hz (125–180). For segmented levels, the amplitude displayed on the programmer differs from this sum whenever fewer or more than three segments are active (e.g. 6 mA displayed for 2.0 mA delivered through the single segment 2c in P04). Contacts, pulse width and frequency were identical at M6 in all patients with an M6 recording, except for the left directional contact of P02 (1c to 1a). Per-contact current differed between the two visits by less than 0.5 mA in three patients (P01, P02, P04), reflecting patient-controlled titration between visits, and was unchanged in the six others.

Supplementary Table 2. DBS stimulation parameters at M3 and M6.

| ID | Visit | Left hemisphere | | | | Right hemisphere | | | |
|---|---|---|---|---|---|---|---|---|---|
| | | Cathode | Intensity (mA/contact) | PW (µs) | Frequency (Hz) | Cathode | Intensity (mA/contact) | PW (µs) | Frequency (Hz) |
| P01 | M3 | 2a/2b/2c | 0.4 | 60 | 125 | 2a/2b/2c | 0.4 | 60 | 125 |
| | M6 | 2a/2b/2c | 0.4 | 60 | 125 | 2a/2b/2c | 0.4 | 60 | 125 |
| P02 | M3 | 1c | 1.0 | 120 | 180 | 2a/2b/2c | 0.3 | 90 | 180 |
| | M6 | 1a | 1.3 | 120 | 180 | 2a/2b/2c | 0.5 | 90 | 180 |
| P03 | M3 | 1a/1b/1c | 0.6 | 40 | 125 | 1a/1b/1c | 0.5 | 60 | 125 |
| | M6 | — | — | — | — | — | — | — | — |
| P04 | M3 | 2c | 2.0 | 70 | 180 | 2a/2b/2c | 0.3 | 60 | 180 |
| | M6 | 2c | 2.3 | 70 | 180 | 2a/2b/2c | 0.5 | 60 | 180 |
| P05 | M3 | 1a/1b/1c | 0.7 | 60 | 125 | 2a/2b/2c | 0.6 | 90 | 125 |
| | M6 | 1a/1b/1c | 0.7 | 60 | 125 | 2a/2b/2c | 0.6 | 90 | 125 |
| P06 | M3 | 2a/2b/2c | 0.7 | 50 | 125 | 1c | 2.0 | 60 | 125 |
| | M6 | 2a/2b/2c | 0.7 | 50 | 125 | 1c | 2.0 | 60 | 125 |
| P07 | M3 | 1a/1b/1c/2a/2b/2c | 0.6 | 60 | 125 | 2a/2b/2c | 0.5 | 60 | 125 |
| | M6 | 1a/1b/1c/2a/2b/2c | 0.6 | 60 | 125 | 2a/2b/2c | 0.5 | 60 | 125 |
| P08 | M3 | 2a/2b/2c | 0.4 | 40 | 125 | 1a/1b/1c | 0.4 | 40 | 125 |
| | M6 | 2a/2b/2c | 0.4 | 40 | 125 | 1a/1b/1c | 0.4 | 40 | 125 |
| P09 | M3 | 1a/1b/1c | 0.7 | 40 | 165 | 2a/2b/2c | 0.6 | 40 | 165 |
| | M6 | 1a/1b/1c | 0.7 | 40 | 165 | 2a/2b/2c | 0.6 | 40 | 165 |
| P10 | M3 | 1a/1b/1c | 0.6 | 40 | 125 | 1a/1b/1c | 0.6 | 40 | 125 |
| | M6 | 1a/1b/1c | 0.6 | 40 | 125 | 1a/1b/1c | 0.6 | 40 | 125 |

Values read from the Percept session reports (TherapySnapshot of the BrainSense streams). Cathode contacts refer to the active contacts of the recording group (Medtronic SenSight B33005 directional lead: ring contacts 0 and 3, segmented levels 1 and 2 with segments a/b/c). All configurations were monopolar (case anode). Intensity is the current per active contact (ElectrodeAmplitudeInMilliAmps); the total programmed amplitude is the sum over the active contacts. PW: pulse width. P03: no M6 recording.

#### S6. Home Event Snapshot mapping and verification

For each patient, home Event Snapshot labels (A–D) were mapped to the corresponding experimental conditions (A: rest/stimulation OFF, B: movement/stimulation OFF, C: rest/stimulation ON, and D: movement/stimulation ON) using a predefined mapping. Stimulation status (ON vs. OFF) was verified directly from the device files. The behavioral condition (rest vs. movement) could not be independently verified and therefore relied on the condition reported by the patient at the time of acquisition. The number of available home Event Snapshots for each patient and condition is reported in Supplementary Table 3.

Supplementary Table 3. Number of available home Event Snapshots by participant and experimental condition. Dashes indicate unavailable home recordings. P03 withdrew after M3; P04 had no home recordings during stimulation OFF.

| ID | Rest OFF | Movement OFF | Rest ON | Movement ON | Total |
|---|---|---|---|---|---|
| P01 | 21 | 22 | 23 | 21 | 87 |
| P02 | 29 | 32 | 26 | 30 | 117 |
| P03 | — | — | — | — | — |
| P04 | — | — | 8 | 7 | 15 |
| P05 | 39 | 39 | 36 | 40 | 154 |
| P06 | 17 | 17 | 16 | 17 | 67 |
| P07 | 15 | 14 | 13 | 13 | 55 |
| P08 | 32 | 34 | 35 | 36 | 137 |
| P09 | 11 | 10 | 12 | 11 | 44 |
| P10 | 15 | 24 | 27 | 28 | 94 |
| **Total** | **179** | **192** | **196** | **203** | **770** |

P04 had no home recordings during stimulation OFF and was therefore excluded from home-based analyses for these conditions, but remained included in M3–M6 comparisons.
P03 withdrew after M3 and had no home recordings.

#### S7. In-clinic recording duration and usable data

Supplementary Table 4. Recorded and usable duration (artifact free) of in-clinic LFP recordings by participant, session, and condition. Values are shown for M3 and M6 across rest and movement during stimulation OFF and ON. Each cell reports usable/recorded duration in seconds.

| | M3 | | | | M6 | | | |
|---|---|---|---|---|---|---|---|---|
| **ID** | **Rest OFF** | **Mvt OFF** | **Rest ON** | **Mvt ON** | **Rest OFF** | **Mvt OFF** | **Rest ON** | **Mvt ON** |
| P01 | 213/213 | 120/120 | 130/133 | 151/151 | 238/246 | 206/211 | 148/154 | 149/154 |
| P02 | 147/147 | 150/150 | 154/154 | 146/146 | 33/33 | 38/38 | 40/40 | 117/117 |
| P03 | 152/152 | 63/74 | 154/154 | 162/170 | — | — | — | — |
| P04 | 178/178 | 62/62 | 164/167 | 61/61 | 82/82 | 29/29 | 74/74 | 74/74 |
| P05 | 44/44 | 38/38 | 48/48 | 36/39 | 65/65 | 62/62 | 65/66 | 60/63 |
| P06 | 42/42 | 42/42 | 36/36 | 35/35 | 150/150 | 32/32 | 26/37 | 34/40 |
| P07 | 34/34 | 35/35 | 34/36 | 44/48 | 71/71 | 28/28 | 38/38 | 43/43 |
| P08 | 125/125 | 124/124 | 117/122 | 122/126 | 66/66 | 54/54 | 52/61 | 54/61 |
| P09 | 78/80 | 82/86 | 114/121 | 117/121 | 89/93 | 53/55 | 58/61 | 58/60 |
| P10 | 41/43 | 57/58 | 70/73 | 74/76 | 46/47 | 50/50 | 61/63 | 58/61 |
| **Mean ±SD** | **105±66** | **77±41** | **102±51** | **95±50** | **93±64** | **61±56** | **62±35** | **72±37** |

Values are usable/recorded duration (s), rounded to the nearest second, where usable duration corresponds to data retained for PSD analysis after quality control. Mean ± SD is of usable duration only. P03 had no M6 recording.

#### S8. Signal processing

##### *Handling of censored spectral values in at-home spectra*

Some at-home spectra contained negative values that remained constant across consecutive high-frequency bins. According to the device specifications, these values may represent censored measurements. This affected the at-home Percept spectra only; the clinical M3/M6 spectra were computed offline and cannot take negative values. Before squaring the spectra or estimating aperiodic components, each negative bin was replaced with the nearest preceding non-negative value along the frequency axis. Because censoring primarily affected the upper-frequency range, gamma power (>35 Hz) was excluded from inferential analyses. The original spectra were retained for visual quality control. The data were also screened for extended plateaus consisting of ≥7 consecutive identical frequency-bin values. Affected recordings and frequency ranges are documented in Supplementary Tables 5 and 6.

Supplementary Table 5. Audit of negative values in home Event Snapshot spectra. Affected participants, recording conditions, frequency ranges, numbers of affected/available segments, and observed negative values are shown.

| ID | Condition | Range (Hz) | Segments affected/total | Value(s) (µVp) |
|---|---|---|---|---|
| P02 | Movement OFF | 58.6–96.7 | 32/32 | -0.111831, -0.111336 |
| P02 | Movement ON | 58.6–96.7 | 30/30 | -0.111831, -0.111336 |
| P02 | Rest OFF | 58.6–96.7 | 29/29 | -0.111831, -0.111336 |
| P02 | Rest ON | 58.6–96.7 | 26/26 | -0.111831, -0.111336 |
| P04 | Movement ON | 58.6–96.7 | 7/7 | -0.111336 |
| P04 | Rest ON | 58.6–96.7 | 8/8 | -0.111336 |
| P09 | Movement OFF | 74.2–96.7 | 10/10 | -0.112330 |
| P09 | Movement ON | 74.2–96.7 | 11/11 | -0.112330 |
| P09 | Rest OFF | 74.2–96.7 | 11/11 | -0.112330 |
| P09 | Rest ON | 74.2–96.7 | 12/12 | -0.112330 |
| **Total** | — | — | **176/176** | — |

Supplementary Table 6. Audit of spectral plateaus in home Event Snapshot recordings. For each participant, the table summarizes affected conditions, proportion of segments containing plateaus, lowest and median plateau onset frequencies, plateau length, and affected frequency bands.

| ID | Conditions affected | Segments with plateau/total | Lowest onset (Hz) | Median onset (IQR), Hz | Median length (IQR), bins | Bands affected |
|---|---|---|---|---|---|---|
| P01 | All | 174/174 | 9.77 | 44.0 (31.2–63.5) | 18 (9–28) | α–γ |
| P02 | All | 234/234 | 12.70 | 45.9 (35.2–58.6) | 14 (10–40) | α–γ |
| P03 | — | — | — | — | — | — |
| P04 | 2/4 | 30/30 | 12.70 | 49.3 (35.2–58.6) | 18 (10–40) | α–γ |
| P05 | All | 308/308 | 9.77 | 42.0 (31.2–61.5) | 16 (9–27) | α–γ |
| P06 | All | 134/134 | 25.39 | 49.8 (37.1–65.4) | 15 (9–24) | β–γ |
| P07 | All | 110/110 | 9.77 | 50.8 (36.1–68.4) | 14 (9–20) | α–γ |
| P08 | All | 274/274 | 12.70 | 54.7 (36.1–67.4) | 15 (9–25) | α–γ |
| P09 | All | 88/88 | 9.77 | 51.8 (36.1–74.2) | 17 (9–24) | α–γ |
| P10 | All | 188/188 | 11.72 | 46.9 (32.2–64.5) | 15 (9–24) | α–γ |

A plateau is a run of ≥7 bit-identical consecutive FFT bins (~0.98 Hz/bin).

##### *Determination of the individual tremor frequency ($f_0$)*

The individual tremor frequency ($f_0$) was estimated from upper-limb accelerometer and surface EMG recordings (Fs = 2000 Hz) acquired during in-clinic M3 and M6 sessions. Signals were band-pass filtered between 2 and 30 Hz (fourth-order zero-phase Butterworth), with EMG first reduced to its burst-amplitude envelope (20–400 Hz band-pass, full-wave rectification); a 2 Hz-wide notch at the stimulation frequency was applied to stimulation-ON recordings, and startup transients and visually identified artifacts were excluded. Power spectral density was computed by Welch's method (2 s Hann windows, 50% overlap, 0.49 Hz resolution). For each limb and timepoint, $f_0$ was the most prominent spectral peak within 2.5–12 Hz in the OFF-stimulation movement condition (falling back on the most prominent available condition, needed once), requiring ≥ 3 dB prominence over the local spectral floor. When the dominant peak was a second or third harmonic, a measured peak within 0.5 Hz of f/2 or f/3, itself above the prominence floor and carrying ≥ 50% of the dominant prominence, $f_0$ was moved onto that measured sub-peak. The accelerometer was the primary source; EMG served as fallback for limbs without a usable accelerometer peak (n=3). Both modalities agreed within 1 Hz on 16 of the 19 limbs carrying both. $f_0$ was estimated independently at M3 and M6 and merged into a single value per participant and per hemisphere: prominence-weighted mean when concordant within 1 Hz, manual arbitration otherwise (P7: M3 retained, no peak below 7 Hz being present at M6). Because stimulation acts on contralateral limb tremor, the $f_0$ of each thalamic electrode is that of the opposite limb. All 20 hemispheres yielded a usable $f_0$ (3.42–5.86 Hz, median 4.27 Hz;interquartile range 3.89–4.88 Hz; range 3.42–5.86 Hz, left–right difference above 1 Hz in one participant), and the analysis bands were defined as $f_0$ ± 1 Hz and $2 \cdot f_0$ ± 1 Hz (Supplementary Table 7).

Supplementary Table 7. Individual tremor frequencies used to define tremor-locked LFP bands. Patient- and hemisphere-specific $f_0$ values were derived from the contralateral tremulous limb; $2f_0$ denotes the second harmonic. Analysis bands were defined as $f_0$ ± 1 Hz and $2f_0$ ± 1 Hz.

| | **Tremor-frequency, $f_0$ (Hz)** | | **Second harmonic, $2f_0$ (Hz)** | |
|---|---|---|---|---|
| **ID** | **L** | **R** | **L** | **R** |
| P01 | 4.39 | 3.91 | 8.78 | 7.82 |
| P02 | 4.88 | 4.88 | 9.76 | 9.76 |
| P03 | 5.86 | 5.86 | 11.72 | 11.72 |
| P04 | 4.88 | 3.91 | 9.76 | 7.82 |
| P05 | 3.83 | 4.39 | 7.66 | 8.78 |
| P06 | 3.42 | 3.42 | 6.84 | 6.84 |
| P07 | 3.42 | 4.88 | 6.84 | 9.76 |
| P08 | 3.66 | 3.91 | 7.32 | 7.82 |
| P09 | 4.15 | 4.39 | 8.30 | 8.78 |
| P10 | 4.88 | 3.91 | 9.76 | 7.82 |
| **Mean ± SD** | **4.34 ± 0.79** | **4.35 ± 0.71** | **8.67 ± 1.58** | **8.69 ± 1.41** |

$f_0$ denotes the patient- and hemisphere-specific tremor frequency derived from the contralateral tremulous limb; $2f_0$ denotes the second harmonic. Analysis bands were defined as $f_0$ ± 1 Hz and $2f_0$ ± 1 Hz.

#### S9. Statistical analysis

##### *Systematic session effects: Linear mixed-effects inference and sensitivity analyses*

Linear mixed-effects models were fitted separately for each feature and recording condition, with timepoint (M3, home and M6) as the fixed effect and participant as a random intercept. Home was the reference level, providing direct estimates of M3–home and M6–home differences; the M3–M6 contrast was obtained from the same fitted model. Models were estimated by restricted maximum likelihood and used all available observations for the corresponding feature and condition.

Statistical inference was based on within-participant permutation testing. Timepoint labels were permuted within participant over 5000 permutations, and two-sided permutation *P* values were calculated for each contrast. Ninety-five per cent confidence intervals were estimated using percentile bootstrap resampling of participants, with 2000 resamples per feature and condition. Because permutation *P* values and bootstrap confidence intervals were obtained using different resampling procedures, minor discrepancies between the two could occur close to the significance threshold.

Standardized effect sizes were calculated as $d = \beta/\sqrt{(\tau^2 + \sigma^2)}$, where $\tau^2$ is the participant-level random-intercept variance and $\sigma^2$ the residual variance. When the random-intercept variance was estimated as zero, the denominator reduced to the residual standard deviation and the standardized effect was interpreted cautiously. For log-transformed outcomes, coefficients were back-transformed where appropriate to aid interpretation.

Benjamini–Hochberg false discovery rate correction was applied separately for each session contrast across the 13 features and four recording conditions within each analysis unit, yielding 52 tests per contrast. FDR-adjusted $P < 0.05$ was considered statistically significant.

##### *Movement-related modulation*

Movement-related modulation was quantified within each participant, session and stimulation state as the paired difference between movement and rest recordings, calculated only when both were available. For log-transformed features, this difference is equivalent to the log ratio of movement to rest.

For each feature, the primary linear mixed-effects model was:

*Diff ~ timepoint + stimulation + (1 | participant)*

with home and stimulation OFF as reference levels. The intercept therefore represented the movement effect at home during stimulation OFF, the timepoint coefficients tested whether the movement effect differed between M3 or M6 and home, and the stimulation coefficient tested whether movement modulation differed between stimulation ON and OFF. Models were estimated by restricted maximum likelihood.

Permutation inference was tailored to each model term. The movement intercept was tested using whole-participant sign flipping of the movement–rest difference, exhaustively enumerating all $2^9 = 512$ possible sign combinations. Timepoint effects were tested by permuting timepoint labels within participant × stimulation strata, and the stimulation effect by permuting stimulation labels within participant × timepoint strata, using 5000 permutations. Ninety-five per cent confidence intervals were obtained from 2000 participant-level percentile bootstrap resamples.

Movement effects within stimulation OFF and ON were additionally estimated from reduced models fitted separately within each stimulation state:

*Diff ~ timepoint + (1 | participant)*

The reported state-specific effect was the equal-weight average of the movement–rest contrast across M3, home and M6. Corresponding *P* values were obtained by whole-participant sign flipping, and confidence intervals by participant-level bootstrap resampling. These state-specific contrasts describe movement modulation within each stimulation state; inference on differences between stimulation states was based on the stimulation term in the primary model.

For features analysed on the natural-log scale, coefficients were back-transformed using exp(β) and expressed as proportional or percentage changes. For features analysed on the log10 scale, coefficients were back-transformed using $10^{\beta}$. Beta peak centre frequency and bandwidth were analysed in Hz and reported as absolute changes.

Benjamini–Hochberg false discovery rate correction was applied separately across the 13 features for each primary model term and separately within stimulation OFF and ON for the state-specific movement contrasts. Left, right and bilateral analyses were corrected independently.

For descriptive visualization, the frequency-resolved movement effect was calculated as log(movement/rest) at each FFT bin, first averaged across available sessions within each participant and then across participants, separately for stimulation OFF and ON. Ninety-five per cent bootstrap confidence bands were obtained by resampling participants. These spectra were used descriptively and were not treated as formal hypothesis tests.

***Electrode impedance assessment***

Monopolar contact impedances were extracted from device telemetry at M3 and M6 for each contact and hemisphere in nine participants. When multiple measurements were available at a given timepoint, values were averaged within participant, hemisphere and contact. Longitudinal changes from M3 to M6 were assessed using paired Wilcoxon signed-rank tests, both for impedance averaged across contacts within each hemisphere and at the individual-contact level. Effect sizes were quantified using matched-pairs rank-biserial correlation, and Benjamini–Hochberg false discovery rate correction was applied separately to the hemisphere-averaged and contact-level analyses. Results are reported in Supplementary Results S11.

#### S10. Software

Raw Percept JSON recordings were converted and organised in MATLAB (The MathWorks Inc.) using the Percept Toolbox[15]. All subsequent analyses were performed in Python 3.11.14 with NumPy 2.4.2, SciPy 1.17.1 and pandas 3.0.1[16–18]. Preprocessing, filtering and epoching used MNE-Python 1.11.0[19]. Spectral parameterisation into aperiodic and periodic components used FOOOF 1.1.1[20]. Linear mixed-effects models were fitted with statsmodels 0.14.6 (MixedLM, restricted maximum likelihood)[21], and Benjamini-Hochberg false-discovery-rate adjustment used its multipletests implementation. Intraclass correlation coefficients and their bootstrap confidence intervals, the permutation tests and the cluster bootstrap were implemented directly in NumPy, the ICCs following Shrout and Fleiss[22] and McGraw & Wong[23]. Figures were produced with Matplotlib 3.10.8[24]. Analysis code is available at https://github.com/sahar-allouch/ecolovim-dbs-lfp-home-vs-clinic.

### 2. Supplementary Results

Supplementary Table 8. Descriptive values of bilateral LFP features across sessions and conditions. Values are median (interquartile range) at M3, home, and M6, separately for rest and movement during stimulation OFF and ON. The final column gives participant numbers.

| Feature | Condition | M3 | Home | M6 | n |
|---|---|---|---|---|---|
| ***Absolute band power*** | | | | | |
| Delta | Rest OFF | 28.5 [22.8, 36.5] | 37 [20.4, 47] | 25.4 [21.9, 35.2] | 9/8/9 |
| | Movement OFF | 29 [27.9, 39.4] | 31.7 [22.8, 57.8] | 33.4 [26.7, 55.7] | 9/8/9 |
| | Rest ON | 23.7 [19.7, 25.8] | 44.4 [20.2, 55.5] | 23.4 [22.6, 73.5] | 9/9/9 |
| | Movement ON | 25.9 [17.8, 36.7] | 42.6 [20.3, 47] | 28.2 [21.5, 119] | 9/9/9 |
| Theta | Rest OFF | 11.5 [8.87, 12.4] | 10.9 [7.07, 16.2] | 10.6 [8.94, 13.2] | 9/8/9 |
| | Movement OFF | 11.1 [9.29, 12.5] | 10.9 [7.46, 15.9] | 11.6 [9.85, 21.1] | 9/8/9 |
| | Rest ON | 9.37 [6.97, 10.7] | 8.52 [6.71, 17.7] | 9.88 [9.21, 16.6] | 9/9/9 |
| | Movement ON | 9.1 [6.75, 9.86] | 7.96 [6.8, 20.5] | 9.44 [8.12, 14] | 9/9/9 |
| Alpha | Rest OFF | 8.77 [6.67, 17.2] | 7 [5.08, 17.9] | 7.35 [7.13, 17.6] | 9/8/9 |
| | Movement OFF | 6.39 [5.58, 7.92] | 7.15 [4.83, 10.4] | 6.27 [5.51, 12.1] | 9/8/9 |
| | Rest ON | 5.65 [5.25, 8.44] | 4.91 [4.36, 11.6] | 6.92 [6.14, 17.5] | 9/9/9 |
| | Movement ON | 4.86 [4.1, 9.73] | 4.91 [3.95, 11.2] | 6.33 [5.98, 12.8] | 9/9/9 |
| Beta | Rest OFF | 22.4 [9.21, 33] | 13.1 [6.7, 25.1] | 15 [13, 29.1] | 9/8/9 |
| | Movement OFF | 18.5 [7.86, 23.3] | 12.1 [6.66, 20.1] | 13.7 [8.93, 23.6] | 9/8/9 |
| | Rest ON | 8.2 [7.32, 15.3] | 7 [4.8, 10.8] | 9.61 [8.74, 20.5] | 9/9/9 |
| | Movement ON | 9.02 [6.66, 16.9] | 6.92 [5.01, 12] | 9.86 [7.26, 20] | 9/9/9 |
| ***Tremor-frequency power*** | | | | | |
| f0 power | Rest OFF | 12.7 [11.8, 18.5] | 14.7 [8.23, 17.6] | 13.2 [9.73, 21.6] | 9/8/9 |
| | Movement OFF | 15.9 [11.9, 17.3] | 16.5 [8.5, 22.5] | 18.5 [13.4, 29.9] | 9/8/9 |
| | Rest ON | 11.2 [9.78, 11.6] | 10.7 [9.56, 29.4] | 13.6 [9.26, 33.4] | 9/9/9 |
| | Movement ON | 13.5 [8.18, 15.3] | 10.5 [10.1, 29.8] | 15.2 [8.04, 30.8] | 9/9/9 |
| 2f0 power | Rest OFF | 6.94 [3.91, 7.79] | 5.2 [3.81, 9.63] | 5.24 [5.03, 7.34] | 9/8/9 |
| | Movement OFF | 5.48 [4.06, 6.07] | 5.17 [3.67, 8.22] | 5.81 [4.5, 9.38] | 9/8/9 |
| | Rest ON | 4.6 [3.98, 5.6] | 4.25 [4.06, 8.41] | 6.16 [4.64, 10.2] | 9/9/9 |
| | Movement ON | 3.88 [3.37, 4.77] | 4.22 [3.43, 8.45] | 5.9 [4.05, 7.58] | 9/9/9 |
| ***Aperiodic features*** | | | | | |
| Exponent | Rest OFF | 1.53 [1.43, 1.56] | 1.69 [1.66, 1.88] | 1.47 [1.37, 1.52] | 9/8/9 |
| | Movement OFF | 1.49 [1.48, 1.54] | 1.71 [1.6, 1.97] | 1.55 [1.39, 1.65] | 9/8/9 |
| | Rest ON | 1.48 [1.42, 1.57] | 1.84 [1.73, 1.97] | 1.57 [1.53, 1.86] | 9/9/9 |
| | Movement ON | 1.53 [1.38, 1.6] | 1.73 [1.71, 1.97] | 1.57 [1.52, 1.78] | 9/9/9 |
| Offset | Rest OFF | 1.57 [1.43, 1.64] | 1.76 [1.46, 1.94] | 1.49 [1.43, 1.78] | 9/8/9 |
| | Movement OFF | 1.52 [1.49, 1.67] | 1.72 [1.5, 2.06] | 1.56 [1.34, 1.89] | 9/8/9 |
| | Rest ON | 1.43 [1.27, 1.5] | 1.76 [1.51, 1.92] | 1.5 [1.45, 1.95] | 9/9/9 |
| | Movement ON | 1.45 [1.32, 1.68] | 1.78 [1.54, 2] | 1.47 [1.46, 1.96] | 9/9/9 |
| Broadband power | Rest OFF | 49 [45, 69.8] | 60.7 [36.8, 72.6] | 52.9 [46.2, 82.6] | 9/8/9 |
| | Movement OFF | 46.8 [44, 64.6] | 56.4 [41.3, 78.8] | 54.8 [43.4, 95] | 9/8/9 |
| | Rest ON | 39 [38.3, 45.2] | 48.1 [32, 68.8] | 58.6 [40.3, 81.2] | 9/9/9 |
| | Movement ON | 44.7 [32.9, 56] | 47.8 [33.6, 75.6] | 54 [39.8, 84] | 9/9/9 |
| ***Beta peak features*** | | | | | |
| Centre frequency | Rest OFF | 22.6 [19.9, 25.1] | 22.1 [20.8, 24.5] | 21.5 [20.4, 25.5] | 9/8/9 |
| | Movement OFF | 23.5 [21.4, 27.2] | 21.8 [20.5, 25.6] | 21.6 [20, 25.2] | 9/8/9 |
| | Rest ON | 19.6 [16.1, 22.4] | 19.1 [16, 21.1] | 18.9 [17.9, 21.6] | 9/9/9 |
| | Movement ON | 20.6 [19.5, 21.6] | 18.3 [15.7, 20.8] | 20.1 [16.8, 24.5] | 9/9/9 |
| Peak power | Rest OFF | 0.512 [0.336, 0.924] | 0.568 [0.237, 0.841] | 0.438 [0.368, 0.62] | 9/8/9 |
| | Movement OFF | 0.652 [0.235, 0.76] | 0.578 [0.195, 0.739] | 0.339 [0.239, 0.571] | 9/8/9 |
| | Rest ON | 0.319 [0.18, 0.358] | 0.337 [0.272, 0.386] | 0.271 [0.237, 0.314] | 9/9/9 |
| | Movement ON | 0.296 [0.157, 0.385] | 0.348 [0.263, 0.363] | 0.227 [0.185, 0.238] | 9/9/9 |
| Bandwidth | Rest OFF | 7.04 [5.55, 8.18] | 6.59 [6.1, 7.01] | 5.04 [4.55, 7.03] | 9/8/9 |
| | Movement OFF | 5.19 [3.69, 6.71] | 6.11 [5.78, 6.63] | 7 [4.5, 7.89] | 9/8/9 |
| | Rest ON | 8.13 [6.56, 9.79] | 8.84 [8, 10.3] | 7.3 [5.96, 9.17] | 9/9/9 |
| | Movement ON | 5.39 [3.72, 8] | 8.96 [7.98, 9.45] | 4.91 [3.45, 8.59] | 9/9/9 |
| Aperiodic-corrected power | Rest OFF | 5.55 [2.86, 7.94] | 5.61 [2.34, 7.9] | 5.56 [4.19, 6.62] | 9/8/9 |
| | Movement OFF | 5.04 [0.978, 7.05] | 5.36 [1.96, 6.97] | 3.45 [2.14, 5.95] | 9/8/9 |
| | Rest ON | 4.14 [1.62, 4.37] | 3.85 [2.65, 4.31] | 3.21 [2.02, 3.95] | 9/9/9 |
| | Movement ON | 2.73 [1.01, 4.06] | 3.42 [2.41, 4.06] | 1.93 [1.5, 3.24] | 9/9/9 |

Values are median [Q1, Q3] across patients within each session (bilateral unit of analysis; one value per patient per session per condition).
n = patients contributing to each session, M3/Home/M6.

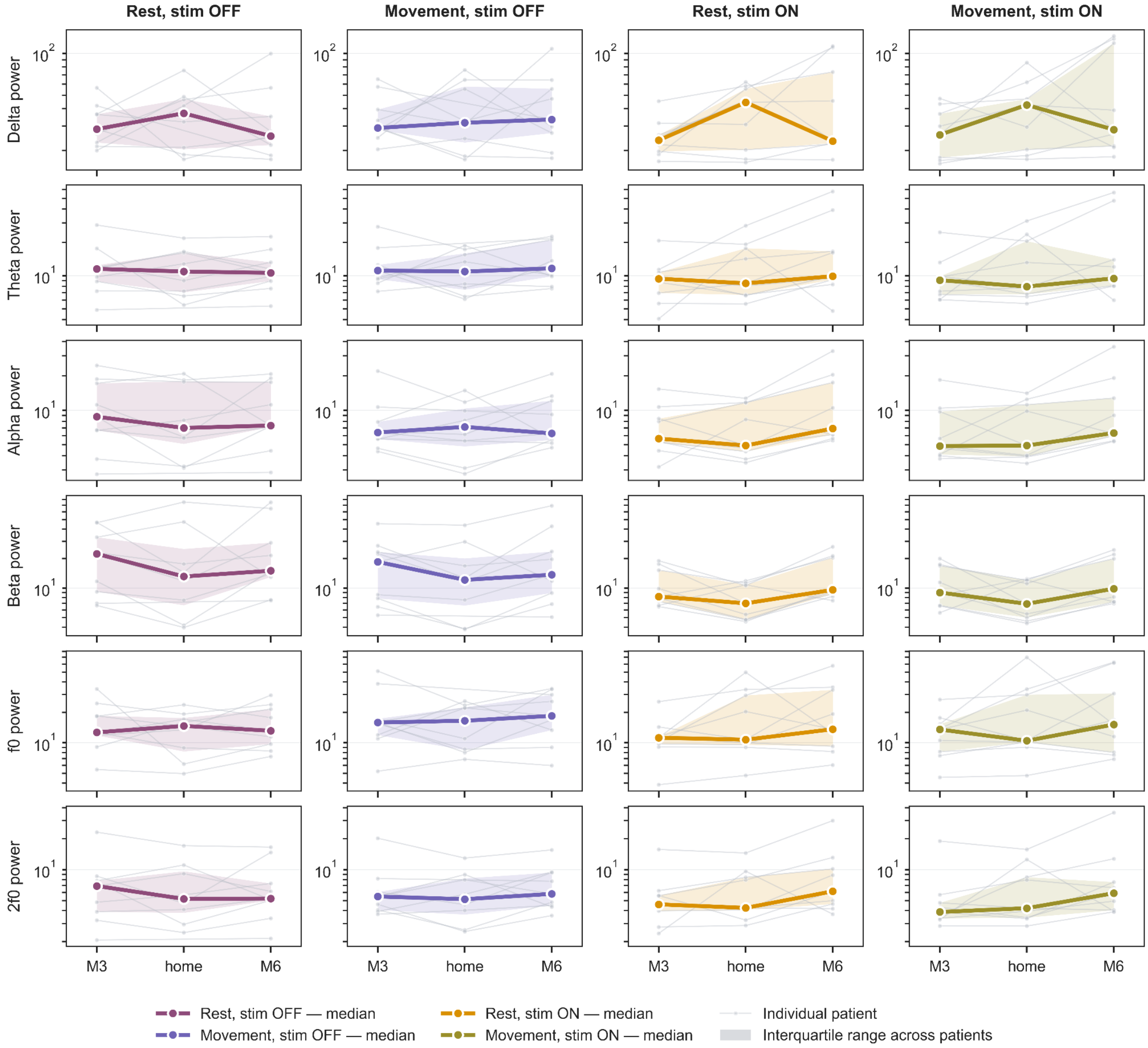


Supplementary Fig. 2. Trajectories of band-limited and tremor-locked LFP power across recording sessions. Participant-level trajectories from M3 to home and M6 are shown for delta, theta, alpha, beta, tremor-frequency ($f_0$), and second-harmonic ($2f_0$) power across rest and movement during stimulation OFF and ON. Thick lines indicate medians, shaded areas the interquartile range, and thin lines individual patients.

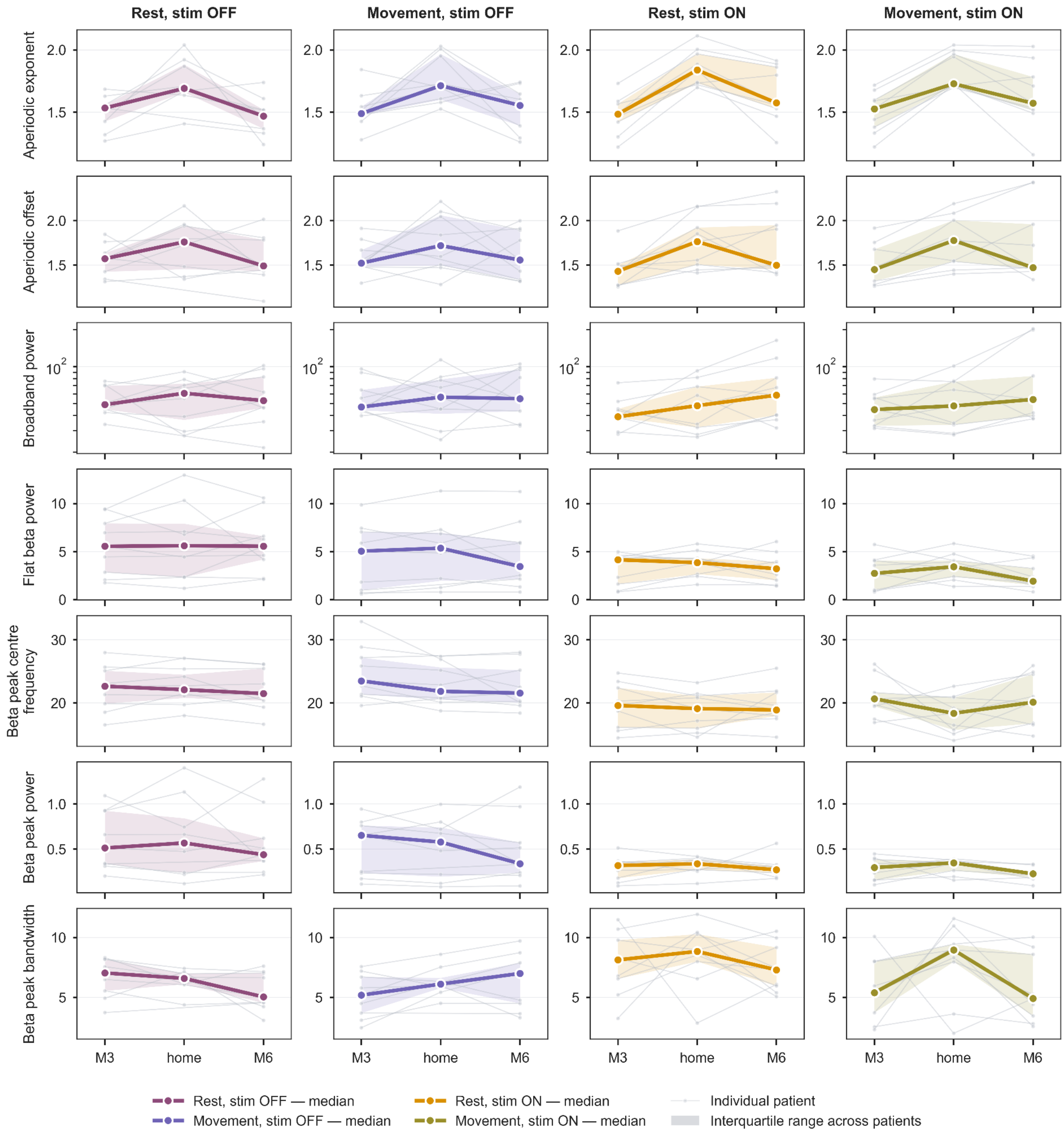


Supplementary Fig. 3. Trajectories of aperiodic and beta-peak LFP features across recording sessions. Participant-level trajectories from M3 to home and M6 are shown for aperiodic exponent, offset, broadband power, aperiodic-corrected beta power, and beta-peak centre frequency, power, and bandwidth across rest and movement during stimulation OFF and ON. Thick lines indicate medians, shaded areas the interquartile range, and thin lines individual patients.

| Feature | Rest OFF | | | Movement OFF | | | Rest ON | | | Movement ON | | |
|---|---|---|---|---|---|---|---|---|---|---|---|---|
| | M3–Home | M3–M6 | Home–M6 | M3–Home | M3–M6 | Home–M6 | M3–Home | M3–M6 | Home–M6 | M3–Home | M3–M6 | Home–M6 |
| **Absolute band power** | | | | | | | | | | | | |
| Delta | -0.26 [-0.96, 0.38] | -0.43 [-0.77, 0.20] | 0.41 [-0.13, 0.79] | -0.04 [-0.52, 0.39] | 0.33 [-0.18, 0.64] | 0.13 [-0.67, 0.68] | 0.34 [-0.05, 0.80] | 0.26 [-0.09, 0.78] | 0.48 [-0.14, 0.86] | 0.50 [-0.09, 0.84] | 0.55 [0.30, 0.68] | 0.39 [-0.19, 0.74] |
| Theta | 0.43 [-0.46, 0.88] | 0.74 [-0.18, 0.96] | 0.70 [0.22, 0.88] | 0.19 [-0.63, 0.62] | 0.67 [-0.10, 0.95] | 0.49 [-0.23, 0.84] | 0.31 [-0.53, 0.88] | 0.54 [0.14, 0.85] | 0.58 [-0.50, 0.82] | 0.48 [-0.20, 0.93] | 0.63 [0.27, 0.88] | 0.61 [-0.48, 0.87] |
| Alpha | 0.79 [0.37, 0.97] | 0.85 [0.31, 0.97] | 0.53 [-0.21, 0.88] | 0.73 [0.40, 0.83] | 0.71 [-0.03, 0.97] | 0.50 [-0.28, 0.75] | 0.63 [-0.16, 0.90] | 0.63 [0.21, 0.76] | 0.66 [0.24, 0.76] | 0.64 [-0.04, 0.95] | 0.62 [0.06, 0.84] | 0.69 [0.21, 0.80] |
| Beta | 0.74 [0.38, 0.90] | 0.87 [0.49, 0.96] | 0.49 [-0.19, 0.81] | 0.80 [0.42, 0.91] | 0.93 [0.75, 0.97] | 0.64 [0.01, 0.84] | 0.34 [-0.18, 0.60] | 0.57 [-0.06, 0.88] | 0.42 [0.05, 0.52] | 0.23 [-0.38, 0.61] | 0.65 [0.10, 0.92] | 0.53 [0.11, 0.65] |
| **Tremor-frequency power** | | | | | | | | | | | | |
| Tremor-frequency power ($f_0$) | 0.25 [-0.73, 0.92] | 0.17 [-0.75, 0.74] | 0.74 [0.19, 0.88] | 0.46 [-0.54, 0.73] | 0.76 [0.15, 0.94] | 0.55 [-0.32, 0.90] | 0.53 [-0.04, 0.92] | 0.48 [0.00, 0.86] | 0.56 [-0.12, 0.89] | 0.60 [0.16, 0.91] | 0.64 [0.27, 0.84] | 0.57 [0.04, 0.84] |
| Second-harmonic power ($2f_0$) | 0.75 [-0.02, 0.94] | 0.87 [0.42, 0.95] | 0.57 [-0.26, 0.89] | 0.72 [0.08, 0.86] | 0.73 [-0.11, 0.94] | 0.60 [-0.17, 0.85] | 0.54 [-0.51, 0.97] | 0.70 [0.20, 0.85] | 0.59 [-0.30, 0.80] | 0.58 [-0.18, 0.96] | 0.68 [-0.10, 0.90] | 0.59 [-0.33, 0.83] |
| **Aperiodic features** | | | | | | | | | | | | |
| Exponent | 0.17 [-0.35, 0.55] | 0.13 [-0.83, 0.70] | -0.04 [-0.40, 0.57] | 0.10 [-0.13, 0.33] | 0.47 [-0.35, 0.76] | -0.01 [-0.64, 0.35] | 0.15 [0.00, 0.27] | 0.53 [0.10, 0.77] | 0.33 [0.00, 0.66] | 0.22 [0.07, 0.30] | 0.64 [0.38, 0.73] | 0.36 [0.05, 0.70] |
| Offset | 0.05 [-0.68, 0.52] | 0.22 [-0.53, 0.68] | 0.20 [-0.37, 0.83] | 0.15 [-0.17, 0.63] | 0.39 [-0.33, 0.79] | 0.10 [-0.61, 0.62] | 0.26 [-0.13, 0.66] | 0.37 [0.04, 0.77] | 0.65 [-0.20, 0.94] | 0.48 [0.07, 0.75] | 0.62 [0.33, 0.73] | 0.62 [-0.22, 0.88] |
| Broadband power | 0.16 [-0.48, 0.72] | 0.41 [-0.30, 0.78] | 0.52 [-0.30, 0.76] | 0.20 [-0.31, 0.60] | 0.08 [-0.67, 0.74] | 0.18 [-0.61, 0.75] | 0.38 [-0.28, 0.82] | 0.33 [-0.03, 0.76] | 0.61 [-0.04, 0.81] | 0.51 [-0.12, 0.86] | 0.55 [0.31, 0.70] | 0.55 [-0.14, 0.76] |
| **Beta peak features** | | | | | | | | | | | | |
| Centre frequency | 0.90 [0.66, 0.97] | 0.94 [0.75, 0.99] | 0.85 [0.27, 0.97] | 0.80 [0.56, 0.94] | 0.37 [-0.14, 0.94] | 0.63 [-0.07, 0.96] | 0.79 [0.45, 0.93] | 0.79 [0.17, 0.95] | 0.59 [-0.23, 0.91] | 0.02 [-0.46, 0.48] | 0.23 [-0.56, 0.76] | 0.33 [-0.18, 0.77] |
| Peak power | 0.83 [0.58, 0.94] | 0.81 [0.18, 0.97] | 0.58 [-0.02, 0.87] | 0.91 [0.45, 0.96] | 0.88 [0.60, 0.94] | 0.78 [0.33, 0.99] | 0.61 [-0.11, 0.89] | 0.43 [-0.09, 0.75] | 0.03 [-0.69, 0.67] | 0.21 [-0.51, 0.67] | 0.35 [-0.08, 0.76] | 0.44 [0.09, 0.70] |
| Bandwidth | 0.34 [-0.36, 0.74] | 0.67 [0.33, 0.83] | 0.17 [-0.25, 0.85] | 0.52 [-0.04, 0.77] | 0.26 [-0.35, 0.65] | 0.62 [-0.03, 0.84] | -0.40 [-1.04, 0.51] | 0.31 [-0.28, 0.78] | 0.22 [-0.41, 0.57] | -0.32 [-0.99, 0.47] | 0.27 [-0.48, 0.84] | 0.34 [-0.03, 0.77] |
| Aperiodic-corrected power | 0.87 [0.65, 0.94] | 0.81 [0.28, 0.97] | 0.66 [-0.01, 0.90] | 0.96 [0.86, 0.99] | 0.93 [0.72, 0.97] | 0.88 [0.45, 0.98] | 0.75 [0.41, 0.86] | 0.64 [0.01, 0.92] | 0.54 [-0.07, 0.79] | 0.46 [0.05, 0.67] | 0.71 [0.39, 0.93] | 0.54 [0.12, 0.79] |

Poor <0.50 Moderate 0.50–0.75 Good 0.75–0.90 Excellent >0.90

between-patient SD = 0 — ICC not estimable

low ICC, error below its upper tertile — ICC uninformative

Supplementary Fig. 4. Pairwise between-session reliability. ICC(2,1) estimates and 95% confidence intervals are shown for M3–home, M3–M6 and home–M6 comparisons across rest/movement and stimulation OFF/ON. Cell shading indicates conventional ICC categories. Solid outlines indicate non-estimable ICCs due to zero between-patient variance; dashed outlines indicate low ICCs considered uninformative because estimation error was below its upper tertile.

| Feature | Rest OFF | | | Movement OFF | | | Rest ON | | | Movement ON | | |
|---|---|---|---|---|---|---|---|---|---|---|---|---|
| | M3−Home | M3−M6 | M6−Home | M3−Home | M3−M6 | M6−Home | M3−Home | M3−M6 | M6−Home | M3−Home | M3−M6 | M6−Home |
| **Absolute band power** | | | | | | | | | | | | |
| Delta | -0.18 [-1.04, +0.80] | -0.01 [-1.09, +0.87] | -0.18 [-1.05, +0.58] | -0.06 [-0.94, +0.77] | -0.22 [-0.96, +0.40] | +0.16 [-0.78, +1.11] | -0.64 [-1.17, -0.13] | ↓ -0.83* [-1.61, -0.18] | +0.19 [-0.53, +0.96] | -0.44 [-0.93, +0.01] | ↓ -0.74* [-1.26, -0.25] | +0.30 [-0.48, +1.12] |
| Theta | +0.24 [-0.35, +0.93] | +0.04 [-0.39, +0.47] | +0.20 [-0.27, +0.62] | +0.13 [-0.70, +0.92] | -0.21 [-0.75, +0.26] | +0.34 [-0.32, +1.01] | -0.35 [-1.00, +0.19] | ↓ -0.74* [-1.33, -0.27] | +0.39 [-0.29, +0.87] | -0.31 [-0.91, +0.16] | -0.59 [-1.11, -0.20] | +0.28 [-0.39, +0.78] |
| Alpha | +0.30 [-0.05, +0.69] | -0.01 [-0.32, +0.34] | +0.30 [-0.29, +0.82] | +0.19 [-0.39, +0.67] | -0.23 [-0.70, +0.16] | +0.41 [-0.36, +0.97] | +0.06 [-0.49, +0.49] | ↓ -0.73** [-1.15, -0.29] | ↑ +0.79** [+0.39, +1.13] | -0.06 [-0.59, +0.42] | ↓ -0.67* [-1.14, -0.22] | ↑ +0.61* [+0.19, +0.98] |
| Beta | +0.40 [-0.07, +0.83] | -0.12 [-0.35, +0.21] | +0.51 [-0.18, +1.10] | +0.39 [+0.02, +0.70] | -0.12 [-0.34, +0.10] | ↑ +0.52* [-0.02, +0.92] | ↑ +0.71* [+0.02, +1.30] | -0.41 [-0.97, +0.16] | ↑ +1.13** [+0.57, +1.56] | +0.65 [-0.15, +1.31] | -0.21 [-0.76, +0.34] | ↑ +0.86* [+0.33, +1.25] |
| **Tremor-frequency power** | | | | | | | | | | | | |
| Tremor-frequency power ($f_0$) | +0.32 [-0.39, +1.26] | +0.01 [-0.77, +0.81] | +0.31 [-0.18, +0.75] | +0.16 [-0.53, +0.81] | -0.14 [-0.62, +0.27] | +0.29 [-0.28, +0.85] | -0.51 [-1.08, -0.03] | -0.65 [-1.25, -0.19] | +0.14 [-0.53, +0.78] | -0.42 [-0.94, +0.03] | -0.53 [-0.99, -0.15] | +0.11 [-0.58, +0.72] |
| Second-harmonic power ($2f_0$) | +0.21 [-0.21, +0.66] | -0.01 [-0.32, +0.31] | +0.21 [-0.32, +0.75] | +0.14 [-0.39, +0.67] | -0.20 [-0.67, +0.19] | +0.34 [-0.25, +0.88] | -0.24 [-0.86, +0.25] | ↓ -0.71* [-1.04, -0.36] | +0.47 [-0.14, +0.95] | -0.35 [-0.91, +0.12] | ↓ -0.65* [-1.07, -0.31] | +0.30 [-0.32, +0.82] |
| **Aperiodic features** | | | | | | | | | | | | |
| Exponent | ↓ -1.45** [-2.23, -0.69] | +0.16 [-0.70, +0.82] | ↓ **-1.61*** [-2.67, -0.75] | ↓ -1.39** [-2.15, -0.61] | -0.03 [-0.66, +0.58] | ↓ -1.36** [-2.36, -0.39] | ↓ **-2.09**** [-2.53, -1.65] | ↓ -0.98** [-1.49, -0.47] | ↓ **-1.11*** [-1.80, -0.53] | ↓ **-1.65**** [-2.01, -1.28] | ↓ -0.66* [-1.14, -0.11] | ↓ -0.99** [-1.65, -0.38] |
| Offset | -0.59 [-1.41, +0.33] | +0.02 [-0.77, +0.66] | -0.62 [-1.58, +0.20] | -0.65 [-1.51, +0.17] | -0.14 [-0.72, +0.44] | -0.51 [-1.52, +0.45] | ↓ -1.05** [-1.63, -0.51] | ↓ -0.98** [-1.65, -0.46] | -0.07 [-0.66, +0.50] | ↓ -0.76* [-1.15, -0.35] | ↓ -0.72* [-1.17, -0.31] | -0.04 [-0.67, +0.58] |
| Broadband power | +0.09 [-0.58, +0.86] | -0.13 [-0.82, +0.48] | +0.22 [-0.51, +0.82] | +0.04 [-0.78, +0.86] | -0.21 [-0.93, +0.56] | +0.25 [-0.74, +1.15] | -0.29 [-0.90, +0.27] | ↓ -0.85* [-1.55, -0.28] | +0.56 [-0.04, +1.10] | -0.16 [-0.69, +0.31] | ↓ -0.73* [-1.27, -0.26] | +0.57 [-0.09, +1.19] |
| **Beta peak features** | | | | | | | | | | | | |
| Centre frequency | -0.22 [-0.45, +0.01] | +0.04 [-0.17, +0.30] | -0.26 [-0.57, +0.02] | +0.42 [+0.10, +0.76] | ↑ +0.58* [+0.02, +1.39] | -0.16 [-0.67, +0.22] | +0.22 [-0.16, +0.64] | -0.02 [-0.43, +0.42] | +0.25 [-0.21, +0.84] | +0.80 [+0.07, +1.56] | +0.11 [-0.68, +1.02] | +0.69 [+0.04, +1.45] |
| Peak power | -0.03 [-0.50, +0.37] | +0.07 [-0.22, +0.47] | -0.10 [-0.81, +0.50] | +0.06 [-0.24, +0.33] | +0.04 [-0.27, +0.37] | +0.02 [-0.45, +0.47] | -0.26 [-0.83, +0.24] | -0.07 [-0.78, +0.67] | -0.19 [-0.86, +0.61] | -0.31 [-1.12, +0.61] | +0.59 [-0.14, +1.32] | ↓ -0.90* [-1.27, -0.47] |
| Bandwidth | +0.30 [-0.45, +0.84] | ↑ +0.77* [+0.36, +1.16] | -0.47 [-1.27, +0.26] | -0.54 [-0.97, -0.07] | -0.62 [-1.41, +0.18] | +0.08 [-0.49, +0.58] | -0.28 [-1.27, +0.88] | +0.15 [-0.53, +0.89] | -0.43 [-1.16, +0.31] | -0.79 [-1.76, +0.31] | +0.00 [-0.76, +0.71] | -0.80 [-1.50, -0.12] |
| Aperiodic-corrected power | -0.06 [-0.45, +0.30] | -0.06 [-0.40, +0.29] | +0.00 [-0.64, +0.54] | -0.09 [-0.29, +0.09] | -0.07 [-0.29, +0.19] | -0.03 [-0.38, +0.27] | -0.40 [-0.78, +0.00] | -0.10 [-0.69, +0.43] | -0.30 [-0.80, +0.27] | -0.43 [-1.08, +0.22] | +0.26 [-0.19, +0.76] | ↓ -0.69* [-1.11, -0.25] |

Significant, survives FDR (p_FDR<0.05)    Significant, nominal p only (p<0.05)    no fill = not significant

↓ / ↑ lower / higher at the first-named session
* p<0.05 ** p<0.01 *** p<0.001, on whichever of p / p_FDR set the fill

Supplementary Fig. 5. Pairwise between-session differences. Standardized LME contrasts (d, 95% CI) are shown across behavioral and stimulation conditions. Shading denotes significant differences before and after FDR correction; arrows indicate lower or higher values at the first-named session.

#### S11. Electrode impedance stability

Electrode impedance remained stable between M3 and M6. Mean impedance showed a modest downward tendency in both hemispheres, from 3654 to 3398 Ω on the left and from 3808 to 3346 Ω on the right, but neither change survived correction for multiple comparisons (left: $P_{FDR} = 0.195$; right: $P_{FDR} = 0.203$).

No individual contact showed a significant change after correction. Fourteen of 16 contacts nevertheless showed lower impedance at M6, indicating a broadly consistent but non-significant downward trend across the cohort. These findings provide no evidence of progressive impedance increase or electrode-related signal degradation that could account for the M3–M6 spectral differences.

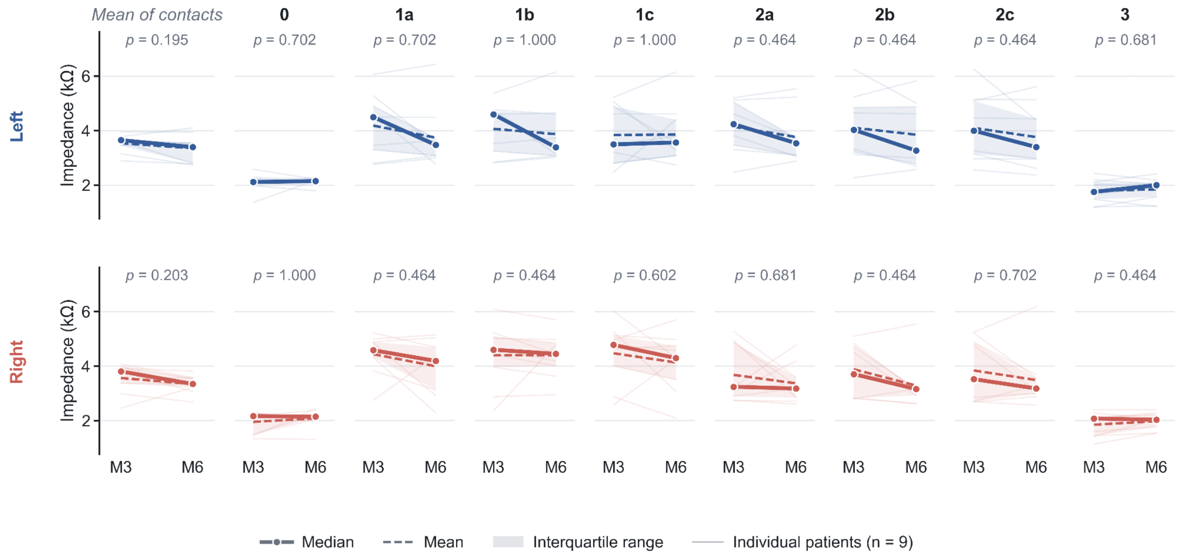


Supplementary Fig. 6. Electrode impedance stability between M3 and M6. Left- and right-hemisphere impedances are shown for the mean across contacts and individual contacts. Thin lines denote individual patients; solid and dashed lines show median and mean, respectively, with interquartile ranges. P values indicate paired M3–M6 comparisons.

### S12. Movement-related modulation

Supplementary Table 9. Movement-related effects within stimulation OFF and ON. Timepoint-averaged movement–rest effects are reported for each spectral feature separately by stimulation state, with β estimates, 95% confidence intervals, nominal p values, and FDR-adjusted p values.

| Feature | Contrast | β [95% CI] | *p* | $p_{FDR}$ |
|---|---|---|---|---|
| **Absolute band power** | | | | |
| Delta | Stim OFF | 0.118 [0.017, 0.225] | 0.090 | 0.195 |
| | Stim ON | 0.074 [-0.008, 0.155] | 0.133 | 0.432 |
| Theta | Stim OFF | 0.120 [-0.077, 0.449] | 0.766 | 0.829 |
| | Stim ON | 0.016 [-0.065, 0.109] | 0.770 | 0.909 |
| Alpha | Stim OFF | -0.143 [-0.345, 0.096] | 0.266 | 0.418 |
| | Stim ON | -0.039 [-0.116, 0.040] | 0.391 | 0.559 |
| Beta | Stim OFF | **-0.198 [-0.293, -0.100]** | 0.004 | **0.025** |
| | Stim ON | -0.002 [-0.060, 0.058] | 0.938 | 0.938 |
| **Tremor-frequency power** | | | | |
| Tremor-frequency power ($f_0$) | Stim OFF | 0.182 [0.057, 0.375] | 0.020 | 0.063 |
| | Stim ON | 0.058 [-0.027, 0.141] | 0.246 | 0.533 |
| Second-harmonic power ($2f_0$) | Stim OFF | 0.035 [-0.191, 0.381] | 0.977 | 0.977 |
| | Stim ON | -0.040 [-0.120, 0.049] | 0.402 | 0.559 |
| **Aperiodic features** | | | | |
| Exponent | Stim OFF | 0.022 [-0.002, 0.060] | 0.152 | 0.283 |
| | Stim ON | -0.002 [-0.024, 0.018] | 0.855 | 0.927 |
| Offset | Stim OFF | 0.040 [-0.019, 0.129] | 0.289 | 0.418 |
| | Stim ON | 0.025 [-0.032, 0.078] | 0.430 | 0.559 |
| Broadband power | Stim OFF | 0.074 [-0.032, 0.248] | 0.371 | 0.482 |
| | Stim ON | 0.069 [-0.005, 0.140] | 0.129 | 0.432 |
| **Beta peak features** | | | | |
| Centre frequency | Stim OFF | **1.097 [0.562, 1.644]** | 0.008 | **0.034** |
| | Stim ON | 0.591 [-0.593, 1.734] | 0.348 | 0.559 |
| Peak power | Stim OFF | -0.087 [-0.143, -0.030] | 0.031 | 0.081 |
| | Stim ON | -0.028 [-0.061, 0.015] | 0.223 | 0.533 |
| Bandwidth | Stim OFF | -0.290 [-1.009, 0.410] | 0.469 | 0.554 |
| | Stim ON | -1.439 [-2.359, -0.478] | 0.035 | 0.305 |
| Aperiodic-corrected power | Stim OFF | **-1.149 [-1.755, -0.597]** | 0.004 | **0.025** |
| | Stim ON | -0.489 [-0.831, -0.109] | 0.047 | 0.305 |

*β* represents the timepoint-averaged movement–rest effect within each stimulation state. 95% CIs are cluster-bootstrap percentile intervals. *p* values are from within-state sign-flip tests; $p_{FDR}$ values were Benjamini–Hochberg corrected across the 13 features within each stimulation state. Bold indicates $p_{FDR} < 0.05$. Units are feature-specific: ln, natural-log ratio; log10, base-10 log ratio; Hz, frequency difference.

Supplementary Table 10. Stimulation-dependent changes in movement-related LFP modulation. Changes in the movement–rest effects between stimulation ON and OFF are reported with β estimates, 95% confidence intervals, nominal p values, and FDR-adjusted p values.

| Feature | Contrast | β [95% CI] | *p* | $p_{FDR}$ |
|---|---|---|---|---|
| **Absolute band power** | | | | |
| Delta | Stim ON vs OFF | -0.046 [-0.157, 0.059] | 0.493 | 0.616 |
| Theta | Stim ON vs OFF | -0.083 [-0.306, 0.063] | 0.255 | 0.414 |
| Alpha | Stim ON vs OFF | 0.118 [-0.062, 0.272] | 0.068 | 0.176 |
| Beta | Stim ON vs OFF | **0.196 [0.117, 0.265]** | <0.001 | **0.003** |
| **Tremor-frequency power** | | | | |
| Tremor-frequency power ($f_0$) | Stim ON vs OFF | -0.129 [-0.246, -0.035] | 0.044 | 0.143 |
| Second-harmonic power ($2f_0$) | Stim ON vs OFF | -0.050 [-0.300, 0.129] | 0.530 | 0.616 |
| **Aperiodic features** | | | | |
| Exponent | Stim ON vs OFF | -0.025 [-0.055, 0.004] | 0.081 | 0.176 |
| Offset | Stim ON vs OFF | -0.018 [-0.087, 0.038] | 0.569 | 0.616 |
| Broadband power | Stim ON vs OFF | -0.004 [-0.140, 0.095] | 0.941 | 0.941 |
| **Beta peak features** | | | | |
| Centre frequency | Stim ON vs OFF | -0.496 [-1.844, 0.748] | 0.509 | 0.616 |
| Peak power | Stim ON vs OFF | 0.059 [0.007, 0.104] | 0.019 | 0.083 |
| Bandwidth | Stim ON vs OFF | -1.165 [-2.392, 0.285] | 0.115 | 0.213 |
| Aperiodic-corrected power | Stim ON vs OFF | **0.649 [0.217, 1.071]** | 0.004 | **0.027** |

*β* represents the change in the movement–rest effect between stimulation ON and OFF. 95% CIs are cluster-bootstrap percentile intervals. *p* values are from permutation tests; $p_{FDR}$ values were Benjamini–Hochberg corrected across the 13 features. Bold indicates $p_{FDR}$ < 0.05. Units are feature-specific: ln, natural-log ratio; log10, base-10 log ratio; Hz, frequency difference.
